# Exome-wide evidence of compound heterozygous effects across common phenotypes in the UK Biobank

**DOI:** 10.1101/2023.06.29.23291992

**Authors:** Frederik H. Lassen, Samvida S. Venkatesh, Nikolas Baya, Wei Zhou, Alex Bloemendal, Benjamin M. Neale, Benedikt M. Kessler, Nicola Whiffin, Cecilia M. Lindgren, Duncan S. Palmer

## Abstract

Exome-sequencing association studies have successfully linked rare protein-coding variation to risk of thousands of diseases. However, the relationship between rare deleterious compound heterozygous (CH) variation and their phenotypic impact has not been fully investigated. Here, we leverage advances in statistical phasing to accurately phase rare variants (MAF ∼ 0.001%) in exome sequencing data from 175,587 UK Biobank (UKBB) participants, which we then systematically annotate to identify putatively deleterious CH coding variation. We show that 6.5% of individuals carry such damaging variants in the CH state, with 90% of variants occurring at MAF < 0.34%. Using a logistic mixed model framework, systematically accounting for relatedness, polygenic risk, nearby common variants, and rare variant burden, we investigate recessive effects in common complex diseases. We find six exome-wide significant (𝑃 < 1.68 × 10^−7^) and 17 nominally significant (𝑃 < 5.25 × 10^−5^) gene-trait associations. Among these, only four would have been identified without accounting for CH variation in the gene. We further incorporate age-at-diagnosis information from primary care electronic health records, to show that genetic phase influences lifetime risk of disease across 20 gene-trait combinations (FDR < 5%). Using a permutation approach, we find evidence for genetic phase contributing to disease susceptibility for a collection of gene-trait pairs, including *FLG*-asthma (𝑃 = 0.00205) and *USH2A*-visual impairment (𝑃 = 0.0084). Taken together, we demonstrate the utility of phasing large-scale genetic sequencing cohorts for robust identification of the phenome-wide consequences of compound heterozygosity.

## Main

Thousands of independent genetic variants have been robustly associated with common, complex human diseases, leading to important advancements in therapeutic development^1^. Naturally occurring variants that disrupt protein-coding sequences are of interest in the context of drug discovery as they modulate potential biological targets with measurable effects on human physiology^2, 3^. Thus, individuals who carry loss-of-function (LoF) variants on both the maternal and paternal copy of a gene, are in principle ‘experiments of nature’ and their identification can help to determine causality between gene function and phenotype^4–6^.

Coding variants in a gene can either be homozygous, when both gene copies harbor the same variant, or CH when both copies harbor different variants, usually at distinct genetic locations within the same gene locus. Alternatively, when two variants are located on a single gene copy, they are said to be ‘in *cis*’. Although both copies of a gene are disrupted in twohit (CH or homozygous) carriers, analyses of the phenotypic impact of coding variation have typically ignored genetic phase information, that is, the separation or ‘phasing’ of an individual’s genome into maternally and paternally derived alleles^7, 8^. Large-scale studies of bi-allelic damaging variation have generally been restricted to homozygotes in populations with excess homozygosity, such as Icelanders^9^, Finns^10, 11^, and consanguineous populations^12^. In contrast, CH are expected to be more common in outbred populations, but are largely under-studied outside of rare disorders^13–17^.

Various methods exist to infer the genetic phase of two variants. ‘Phasing by transmission’ employs family member genotyping and Mendelian inheritance principles^18^, while ‘read-backed phasing’ utilizes physical relationships among variants within sequencing reads^19^. In large-scale biobanks, extensively genotyping family members is impractical, and short-read sequencing technologies only allow read-backed phasing for variants in close proximity. Therefore, ‘statistical phasing’, which models the generative process of newly arising genetic data subject to recombination and mutation^18, 20–23^, is typically used to phase haplotypes in genetic biobank data. Obtaining high-quality statistically phased genetic data requires large sample sizes, (10^5^-10^6^ individuals), and tends to require large reference panels^21^. Furthermore, statistical phasing is more error prone for rare variants, which are precisely the collection of variants that we would like to investigate as they are *a priori* more likely to be deleterious variants of large effect under purifying selection. This difficulty in the accurate statistical phasing of rare variation has historically deterred the analysis of CH variants in biobanks. However, recent advancements in statistical phasing^24^, achieved by combining common variation across genotyping arrays and exome sequencing to create haplotype ‘scaffolds’^22^, enables accurate phasing of rare variants. By using this new accurate phase information which extends down into rare allele frequencies, we can identify damaging CH variants to expand the pool of identifiable two-hit carriers and screen for phenotypic consequences.

We describe and apply a systematic analytical approach to test for autosomal bi-allelic effects, gene-by-gene, across 311 traits in the UKBB 200k exome sequencing (ES) release, combining both CH and homozygous variation. We iteratively refine the candidate associations by adjusting for polygenic background, nearby common variant risk, and rare variant burden within the analyzed gene. Our approach identifies both known and novel bi-allelic-trait associations, providing important insights into the phenotypic impact of gene knockout in humans.

## Results

### Accurate phase inference and validation using parent-offspring trios and short-read sequences

We identified 13,377,336 high-quality variants in 176,935 individuals exome sequenced in the UKBB (Methods). To identify variants co-occurring on the same haplotype (in *cis*) or on opposite haplotypes (in *trans*) gene-by-gene, we jointly phased ES and genotype array data in the UKBB using SHAPEIT5^25^ (Methods) following an investigation into the performance of current phasing software (Supplementary Table 4). Rare variants (MAF < 0.001) are assigned a posterior probability (PP) of true haplotype assignment, known as the phasing confidence score. Confidence in our ability to accurately statistically phase variants decreases with MAF (Supplementary Fig. 5). However, we *a priori* expect a disproportionate recessive damaging signal to reside in CH variants with at least one rare variant, and as a result, choosing a PP cutoff represents a trade-off in the signal to noise ratio. Following phasing, we restrict to 176,587 individuals of genetically-ascertained non-Finnish European (NFE) ancestry (Methods).

To assess statistical phasing quality, we benchmarked against phasing determined with parentoffspring trio data and read-backed phasing. We quantified phasing quality before and after filtering by PP ≥ 0.9 in 96 parent-offspring trios by calculating switch error rates (SER), estimated using Mendelian transmission, across 2,044,234 unique variants stratified by minor allele count (MAC) (Fig. 1a, Supplementary Fig. 6, Supplementary Tables 5-6). Across the 96 children, 93.1% of protein coding genes contained variants that were phased without switch errors (Supplementary Table 7). SERs among singletons (MAC=1) and variants with 2 ≤ MAC ≤ 5 were 12.1% (95% CI = 8.42 − 17.2) and 0.27% (0.13–0.53), respectively (Fig. 1a). Although calculation of SER using trios is the gold-standard approach for phasing quality estimation^21^, it is limited by the number of parent-offspring trios available. For this reason, we also performed read-backed phasing of 62,762 unique pairs of variants using UKBB short read sequences on chromosomes 20-22 in 176,586 NFE individuals using WhatsHap^26^ (Methods). While read-backed phasing only permits ascertainment of genetic phase among pairs of variants spanning one or a few overlapping short read sequences (with typical lengths of 150-300 bp), read-backed phasing accuracy is independent of allele frequency, and therefore represents an orthogonal approach for evaluating the quality of statistically phased variation. Consistent with trio-SER, we observed increasing agreement between pairs of statistically and read-backed phased variants with increasing MAC (Supplementary Fig. 7, Supplementary Table 8). Filtering to phased variants with PP ≥ 0.9, singletons and variants with 2 ≤ MAC ≤ 5, agreement between read-backed phasing and statistical phasing was 85.1% (95% CI = 83.7 − 86.3%) and 99.1% (95% CI = 98.98 − 99.16%) respectively (Supplementary Table 8, Supplementary Fig. 8). Taken together, our benchmarking suggests that statistical phasing of the UKBB dataset is of high quality for rare to ultra-rare variants, increasing our confidence in the identification of damaging CH variation. Given our observations of well-calibrated PP, and the distribution of phasing confidence binned by MAC, we selected the empirical cutoff of PP ≥ 0.9 to retain 8,616,236 variants (43% of which are singletons) for downstream characterization and testing (Supplementary Fig. 5).

**Fig. 1:**
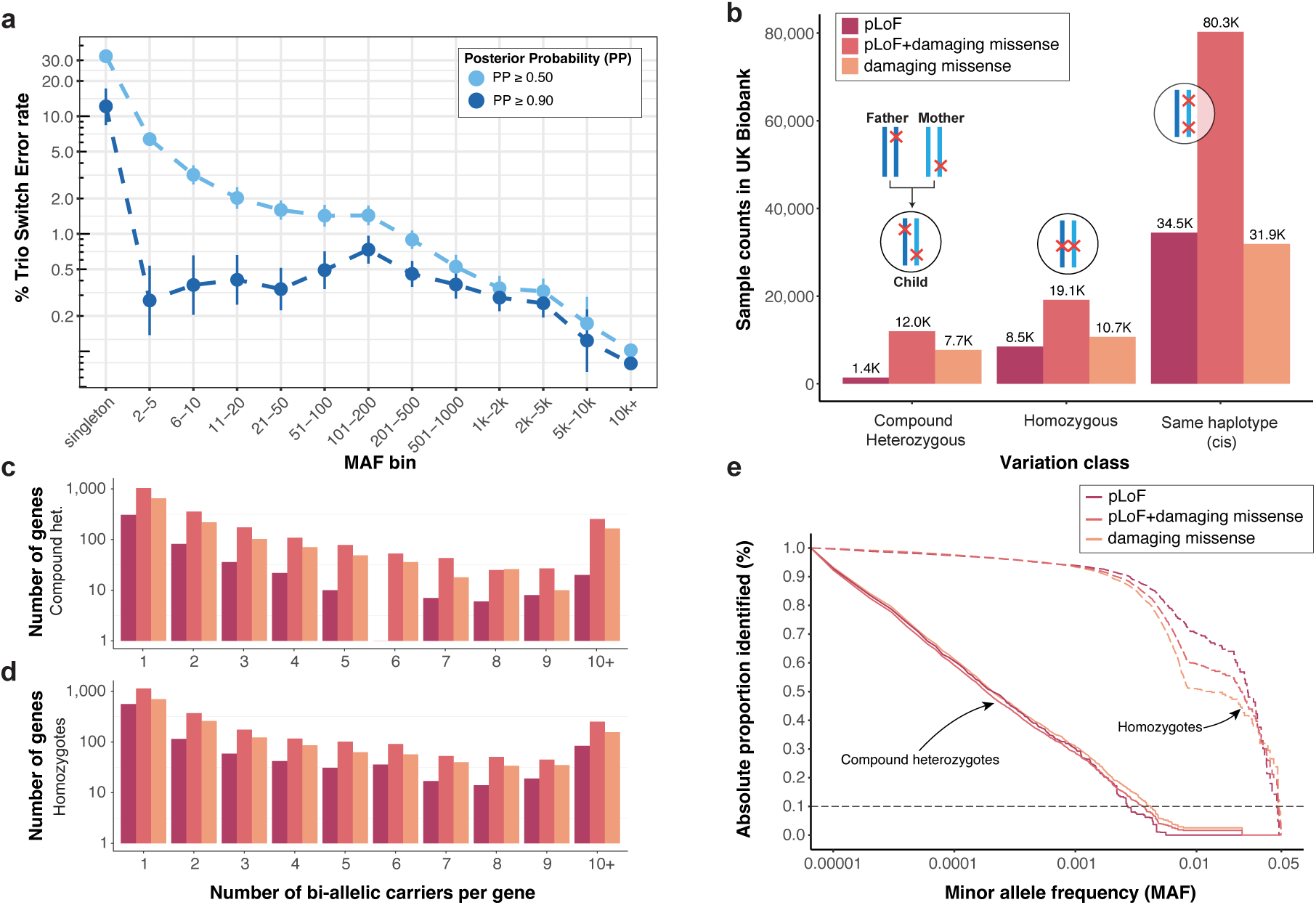
CH variants composed of at least one ultra-rare variant (MAC ≤ 10) can be robustly identified in large scale biobanks. a) Trio SER depicted on 𝑦-axis as a function of MAC bin (𝑥-axis) for phased variants with MAF ≤ 5%, stratified by phasing confidence score PP ≥ 0.5 or PP ≥ 0.9. b) Counts of samples harboring different classes of variation with at least two variants in UKBB. Each set of three bars depicts the number of individuals with at least one CH variant, homozygous variant, or multi-hit (*cis*) variant, respectively. Here, we define a CH pLoF+damaging missense variant as any combination of pLoF and/or damaging missense variation on opposite haplotypes. A qualifying carrier for each bar occurs according to the configuration displayed above the bars, and is grouped by variant consequence according to the color legend. c-d) Number of CH or homozygous carriers per gene. e) 1 - cumulative fraction (𝑦-axis) of homozygous (dashed line) and CH carriers as a function of lowest MAF (𝑥-axis) in bi-allelic variant pairs for which both variants phased at PP ≥ 0.9 (solid line), stratified by variant consequence according to the color key.

### Identification and examination of CH variation in the UKBB

To interrogate the functional role of mono- and bi-allelic variation in the population, we annotated 8,616,236 variants with PP ≥ 0.9 and MAF ≤ 5% across 17,998 autosomal protein-coding genes. We enriched our search for variants with putatively large effect sizes by restricting analyses to two categories of predicted damaging variation: first, we annotated 289,981 high-confidence protein truncating variants, including stop-gain, essential splice and frame-shift variants identified as high-confidence by Loss-Of-Function Transcript Effect Estimator (LOFTEE)^27^, which we refer to as ‘putative loss-of-function (pLoF) variants’. Second, we annotated 444,804 missense variants classified as damaging by both REVEL score ≥ 0.6 and Phred scaled Combined Annotation Dependent Depletion (CADD) score ≥ 20, or LOFTEE low confidence (LC) protein truncating variants; we refer to these variants collectively as ‘damaging missense/protein altering’ (Supplementary Fig. 9, Supplementary Table 9). For each individual, we then determined the set of genes predicted to be affected by pLoFs+damaging missense/protein-altering variants in a CH, homozygous or in *cis* state on the same haplotype.

As we *a priori* expected that essential genes would be less permissible to bi-allelic damaging variants when compared to non-essential genes, we investigated tolerance towards predicted bi-allelic pLoF and pLoF+damaging missense/protein-altering variants across the genome. As some genes carry bi-allelic variants more often than others (owed to a variety of factors such as gene length and baseline mutation frequency^28^), we fit counts of the number of individuals carrying bi-allelic variants per gene using a Poisson regression model accounting for variation in gene length and mutation rate (Methods, Supplementary Tables 10-11). Both pLoF and pLoF+damaging missense/protein-altering bi-allelic variants (homozygous and CH) were significantly depleted in five of the six analyzed essential gene-sets 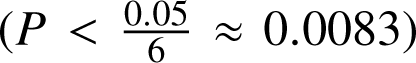 (Supplementary Fig. 12). Conversely, across three non-essential gene-sets, bi-allelic pLoFs+damaging missense/protein-altering variants were enriched among LoF tolerant genes^27^ 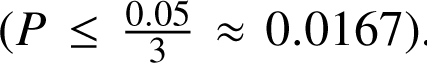. We found that the degree and direction of effects were consistent across CH, homozygous bi-allelic, and heterozygous variants (Supplementary Fig. 12).

In founder^9^ and bottle-necked^10^ populations, some alleles are enriched to high frequency by chance, resulting in better powered association studies for the subset of variant alleles that are inherited from the parental population^6^ at higher frequency. To explore the diversity of bi-allelic variation in UKBB, a largely outbred population, we enumerated two-hit carriers across 176,587 individuals. We observed complete bi-allelic ‘knockout’ of 1,174 unique genes strictly owed to pLoF variants, identifying 1,431 (0.8%) CH and 8,582 (4.8%) homozygous individuals with bi-allelic pLoF variants in at least one gene (Fig. 1b). Across genes, 307 (26.1%) CH and 560 (47.7%) homozygous ‘knockouts’ were observed only in a single individual (Fig. 1c-d). We reasoned that inclusion of damaging missense/protein-altering variants in addition to pLoFs, would expand the number of identifiable damaging bi-allelic variants compared to assessing the two categories independently. Across 3,288 unique genes, we observed 11,491 (6.5%) CH and 17,863 (10.1%) homozygous carriers of pLoF+damaging missense/protein-altering variants. Of these, 1,112 (0.6%) CH and 436 (0.2%) homozygotes were carriers of bi-allelic pLoF+damaging missense/protein-altering variants in genes linked to traits with autosomal recessive mode of inheritance in Online Mendelian Inheritance in Man (OMIM)^29^. We generally observed a higher prevalence of carriers with variants in *cis* compared to CH, with over a third of individuals (64,555, 36.6%) carrying ≥2 pLoF+damaging missense/protein-altering variants on a single haplotype (Fig. 1b).

To better understand the evolutionary dynamics giving rise to pathogenic variants in *trans*, we examined the spectrum of allele frequencies of the constituent variants among our confidently called damaging CHs variants. CHs variants tend to comprise of two variants where one resides on a common haplotype, while the other on a rare haplotype, with a median difference in MAC of 1,181 (Supplementary Fig. 13-14). Approximately 90% of CH-constituent variants have MAF ≤ 0.0038, compared to homozygotes in which 90% are detected at MAF ≥ 0.0022 (Fig. 1d), suggesting that identifying deleterious bi-allelic CH variants requires phasing of rare alleles (Supplementary Fig. 15-16).

Multiple studies have assessed the prospects of ascertaining bi-allelic LoF variation at larger sample sizes in consanguineous, bottle-necked, and outbred populations^6, 12^. To investigate empirically how the number of unique genes with bi-allelic variants scales in an outbred population, we performed down-sampling of UKBB participants. Consistent with previous literature, additional CH and homozygous variants can be inferred by considering both pLoF and damaging missense/protein-altering variation at even larger sample sizes (Supplementary Fig. 19).

### Systematic evaluation of bi-allelic effects on common disease

We performed a series of association analyses using Scalable and Accurate Implementation of GEneralized mixed model (SAIGE)^30^, a generalized mixed model that uses a saddle-point approximation to provide accurate 𝑃-values for traits with extreme case-control ratio imbalance. This allowed us to investigate the effects of bi-allelic variants in 176,587 individuals across 311 phenotypes with varying population prevalence identified from primary and secondary care electronic health records (EHRs) (Methods). We restricted to 952 protein-coding genes with at least 5 individuals carrying bi-allelic variants in the same gene, which allowed us to detect odds ratio (OR) ≥ 10, for traits at approximately 2% population prevalence, with 80% power at exome-wide significance (Bonferroni 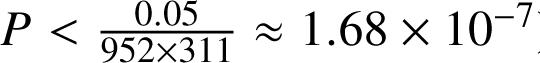) (Methods, Supplementary Fig. 20). Using simulations, we confirmed our ability to detect recessive signals of association with well-calibrated false positive rates across a range of effect sizes (Methods, Supplementary Fig. 21a-c). We tested a total of 299,854 gene-trait combinations, and identified 30 gene-trait associations at nominal significance 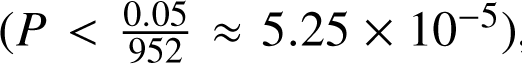, of which seven remained significant following stringent Bonferroni correction (𝑃 < 1.68 × 10^−7^) (Supplementary Table 14, Supplementary Fig. 10).

A recessive gene-trait association may be driven by a variety of genetic factors unrelated to CH or homozygous status, such as polygenic background or through genetic tagging of a nearby common variant association. To mitigate these factors, we created a pipeline to condition on external genetic effects within the gene-trait regression model (Methods). First, we trained polygenic risk scores (PRS) for 111 significantly heritable traits (*h*^2^_snp_ 𝑃 < 0.05 and 𝑛_eff_ ≥ 5000) using LDPred2^31^ (Methods, Supplementary Table 12), a tool that allows PRS derivation based on summary statistics and linkage information. To control for polygenic risk and potentially boost power for association^32^, we included the off-chromosome PRS as an additional covariate (Supplementary Table 14). While the resulting *P*-values were altered by less than a single order of magnitude with the incorporation of PRS (Supplementary Fig. 11), controlling for PRS resulted in the abrogation of four nominally significant (𝑃 < 5.25 × 10^−5^) gene-trait associations. To capture the effects of any causal common variants in linkage disequilibrium (LD) with the pLoF or damaging missense/protein-altering variants constituting the CH or homozygous variant, we further conditioned on nearby (within 1 mega base pairs (Mb) of the associated gene) common (MAF > 1%) variant association signals (Methods, Supplementary Table 13), which abrogated (𝑃 > 0.05) the signal of two gene-trait pairs.

Lastly, we investigated whether any of the identified, putative recessive, associations could be accounted for by assuming an additive genetic architecture. To do this, we counted the number of gene copies affected by pLoF+damaging missense/protein altering variants in each individual. For each putative recessive association, we re-ran the analysis while simultaneously conditioning on the number of affected haplotypes. We also employed a complementary variant-level approach and repeated the analysis, conditioned on all low-frequency (MAC > 10, MAF < 5%) and ultra-rare (MAC ≤ 10) damaging variants (pLoF+damaging missense/protein-altering), including those that constitute the bi-allelic variant in question. Conditioning on the additive effects abrogated the signal of a single nominally significant gene-trait pair (𝑃 < 5.25 × 10^−5^) (Supplementary Table 14).

Together, these analyses refined the list of putative gene-trait associations to 23 nominally significant associations out of which six are significant after correcting for multiple testing (conservative Bonferroni 𝑃 < 1.68 × 10^−7^) (Fig. 2a-2b, Supplementary Table 14) comprising 17 unique genes and 22 traits. Notably, only six of the 23 associations remained nominally significant (𝑃 < 5.25 × 10^−5^) when restricted to only CH variant-carriers, and just four of 23 remained nominally significant when testing homozygous variants alone, underscoring the power of jointly analyzing these variant sets.

**Fig. 2:**
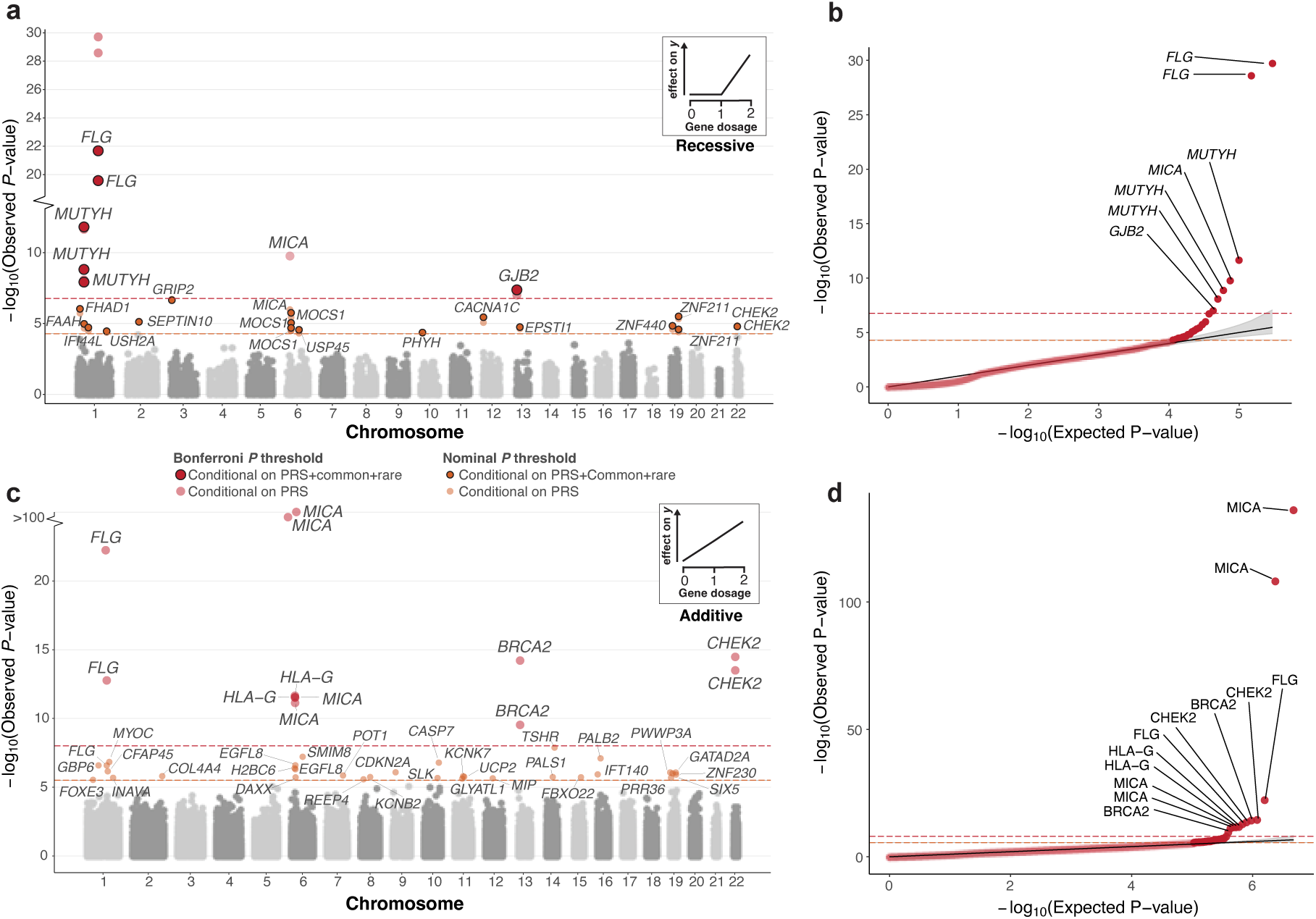
Conditional recessive and additive modeling of gene copy disruption in 311 pheno-types across 176,587 participants. a) Recessive Manhattan plot depicting log_10_-transformed gene-trait association *P*-values against chromosomal location. Associations are colored red or orange based on whether they are Bonferroni (𝑃 < 1.68 × 10^−7^) or nominally (𝑃 < 5.25 × 10^−5^) significant. Transparent coloring represents the resulting *P*-value when conditioning only on PRS, whereas solid coloring with black outline represents the *P*-value derived after conditioning on off-chromosome PRS, nearby (500 kb) common variant association signal, and rare variants within the gene when applicable (methods). The Bonferroni and nominal significance thresholds are also displayed as orange and red dashed lines respectively. A gene may appear multiple times if it is associated with more than one phenotype. A qualifying example of the recessive inheritance pattern is shown in the top right: disruption of both gene copies result in an effect on the phenotype (*y*). b) QQ-plot for genes with bi-allelic damaging variants after conditioning on off chromosome PRS. The shaded area depicts the 95%CI under the null. Gene-trait associations passing Bonferroni significance are labeled accordingly. c-d) Additive Manhattan plot and corresponding QQ-plot for genes with mono and bi-allelic damaging variants associated with at least one phenotype after conditioning on off chromosome PRS when applicable (methods). No additional conditioning was performed in this analysis. Gene-trait associations are colored red and orange based on whether they are respectively Bonferroni (𝑃 < 9.8 × 10^−9^) or nominally (𝑃 < 3.05 × 10^−6^) significant. The additive inheritance model is depicted in the top right: each affected haplotype result in a incremental effect on the phenotype (*y*).

We observed recessive gene-trait relationships across multiple organ systems (nervous, respiratory, circulatory, and genitourinary among others). All six associations that met the significance threshold after Bonferroni correction (𝑃 < 1.68 × 10^−7^) have previously been reported in the literature. For example, individuals with bi-allelic variants in *MUTYH*, a gene involved in oxidative DNA-damage repair^33^, are at significantly increased risk of developing colorectal cancer (log_10_(OR) = 4.7 (95% CI = 3.38 − 6.01), 𝑃 = 2.2 × 10^−12^). We also find that bi-allelic variants in *FLG* increase risk of both asthma^34^ (log_10_(OR) = 0.33 (0.26 − 0.39), 𝑃 = 2.09 × 10^−22^) and dermatitis^35^ (log_10_(OR) = 0.28 (0.22 − 0.33), 𝑃 = 2.65 × 10^−20^). In addition, we observe that bi-allelic variants in *GJB2* increase the risk of hearing loss^29^ (log_10_(OR) = 1.66 (1.05 − 2.26), 𝑃 = 9.93 × 10^−8^). At nominal significance (𝑃 < 5.25 × 10^−5^), 10 of 23 associations have previously been reported. For example, bi-allelic variants in *USH2A*, linked to retina homeostasis^36^, increase risk of visual impairment (log_10_(OR) = 5.77 (2.93 − 8.62), 𝑃 = 3.5 × 10^−5^). For the remaining unreported hits, we observe gene-trait associations with plausible mechanistic insights. For example, we observe that putatively damaging bi-allelic variation in FAAH, a fatty acid amide hydrolase^37^, are associated with increased risk of dementia (log_10_(OR) = 22.92 (12.35−33.48), 𝑃 = 1.06×10^−5^), consequently offering evidence supporting the hypothesis that lipid metabolism dysfunction is central to dementia pathogenesis^38^.

### Boosting power in gene-level regression models through rare variant haplotype collapsing

Complementary to the recessive models described above, rare variant burden testing, which involves the aggregation of rare variants within a gene, has proven to be a robust method to collectively assess the phenotypic impact of rare variation across individuals. Rare variants are aggregated due to their low allele frequency leading to lack of statistical power for detection of single-marker associations. However, these frameworks generally ignore the genetic phase within each individual, and therefore do not differentiate between scenarios in which multiple damaging variants reside on the same (*in cis*) or opposite (*in trans*) haplotypes, despite these two forms having potentially distinct functional and phenotypic effects. We conducted additive genome-wide association analyses by testing for associations between the the number of disrupted gene copies (across 16,363 protein-coding genes with at least 10 haplotypes carrying pLoF+damaging missense/protein altering variation) in an individual and case status (across 311 phenotypes) (Methods, Fig. 2c-2d). After adjusting for polygenic contribution, we found 38 nominally significant gene-trait associations (Nominal 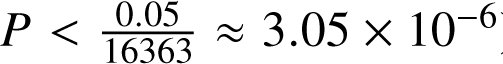), among which 12 were significant associations after multiple-testing correction (Bonferroni 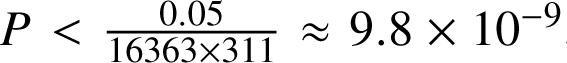, Supplementary Table 15). Among the significant hits are previously reported associations, including association between the number of putatively damaged copies of *BRCA2* (𝑃 = 6.16 × 10^−15^) and *CHEK2* (𝑃 = 3.34 × 10^−15^), and breast cancer.

### Permutation testing to establish the impact of genetic phase on disease risk

It is commonly accepted that compound heterozygosity drives recessive disease risk by disruption of both copies of an implicated gene^13–15^. However, this notion has not been well studied in a large-scale population cohort. To assess the degree to which compound heterozygosity, rather than co-occurring variants on the same haplotype, drives disease risk, we permuted the genetic phase of observed pLoF+damaging missense/protein-altering variants within a gene to generate an empirical distribution of 𝑡-statistics corresponding to disease-association strength in the absence of phase information (Fig. 3a-b). To ensure a sufficiently large sampling distribution, we restricted our analysis to 5 nominally significant (𝑃 < 5.25 × 10^−5^) gene-trait combinations with at least ten individuals that are either CH variant-carriers or with two or more pLoF or damaging missense/protein-altering variants on the same haplotype (Methods).

**Fig. 3:**
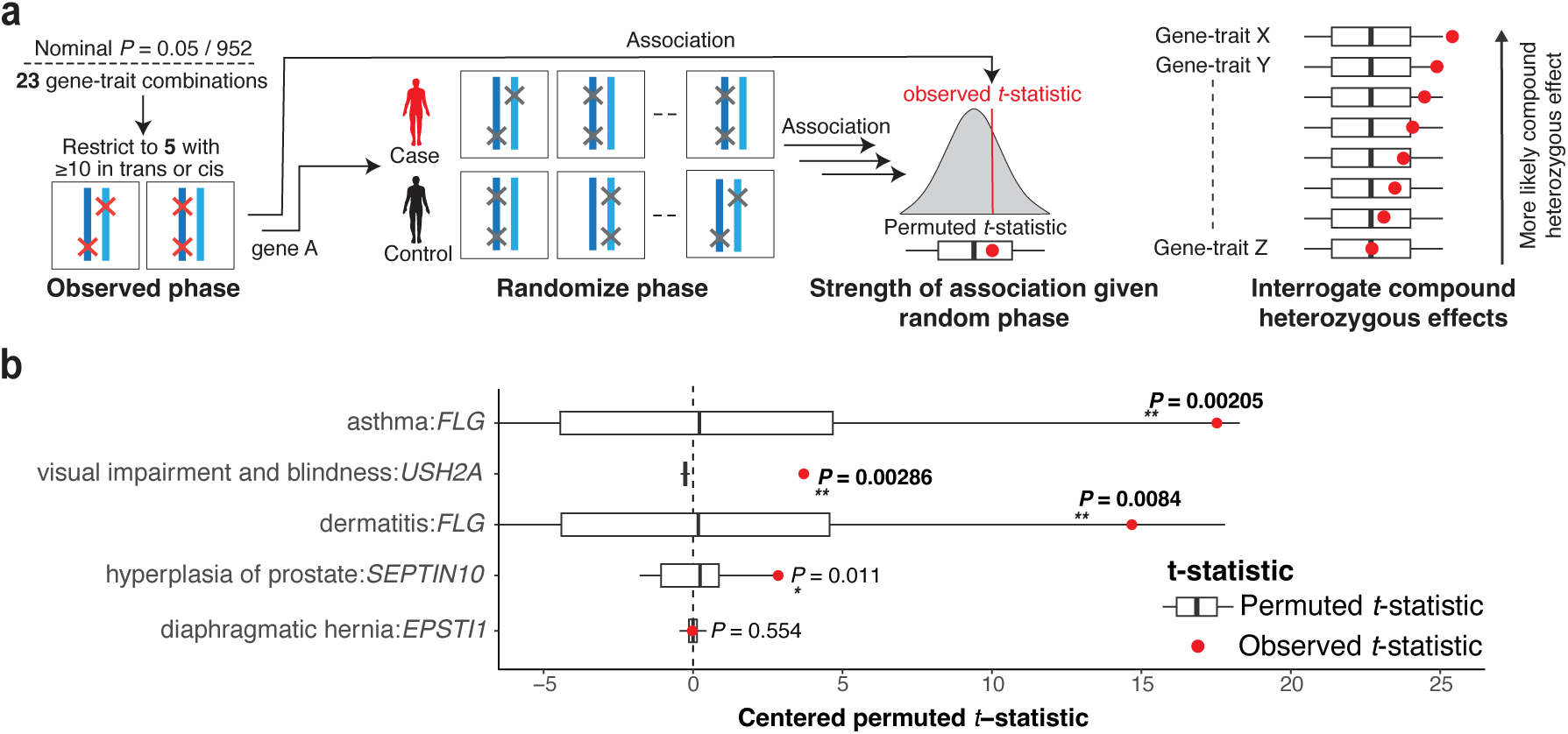
*In-silico* permutation of genetic phase provides evidence for CH-specific effects. a) Overview of the permutation pipeline. To be sufficiently powered to detect effects, we considered five significant (𝑃 < 0.01) gene-trait pairs from the genome-wide analysis that have at least ten individuals harboring pLoF or damaging missense/protein-altering variants on the same haplotypes or CH carriers. Then, we shuffled CH *trans* and *cis* labels across samples and re-ran the association analysis, resulting in a null distribution of permuted 𝑡-statistics corresponding to the association strength in the absence of phase information. We derive the one-tailed empirical 𝑃-value by comparing the observed 𝑡-statistics with the empirical null distribution. b) The resulting distributions of permuted (white and black box plots) and observed 𝑡-statistic (red dot) for each gene-trait and the resulting empirical 𝑃-value. 𝑃-values shown in bold indicate Bonferroni significance (𝑃 < 0.05/05 = 0.01). Box and whisker plots display the quartiles of the empirical null distribution.

We found evidence for significant (Bonferroni 𝑃 = 0.05/5 = 0.01) compound-heterozygous specific effects in three of the five analyzed gene-trait combinations: CH variants in *FLG* are associated with increased risk of both asthma (𝑃 = 0.00205) and dermatitis (𝑃 = 0.0084), while CH variants in *USH2A* are associated with increased risk of visual impairment and blindness (𝑃 = 0.00286) (Fig. 3c). We identified an additional gene-trait association at nominal significance (𝑃 < 0.05), namely CH variants in *SEPTIN10* associated with hyperplasia of prostate (𝑃 = 0.011). Of these, *FLG*-asthma, *FLG*-dermatitis, and *USH2A*-visual impairment associations have previously been linked to disease in the CH state^39–41^. These observations demonstrate, on a large scale, the effect of compound heterozygosity in driving disease susceptibility, and by extension, how appropriately integrating genetic phase can lead to increased power to discover gene-trait associations.

### Non-additive effects of compound heterozygous variants elevate lifetime risk of disease

Bi-allelic effects may be associated with earlier age at onset of disease, which is also often correlated with disease severity. We therefore explored whether CH and homozygous variants had longitudinal effects by evaluating age-at-diagnosis of 278 phenotypes with Cox proportional-hazards models. To identify effects owed to disruption of both gene copies, as opposed to haploinsufficiency, we compared bi-allelic variant carriers against a reference group comprising carriers of a single heterozygous variant for each gene. We tested 267,400 gene-trait combinations with at least five bi-allelic variants (homozygotes or CH) and 100 heterozygotes (Fig. 4a). After adjustment for polygenic risk via off-chromosome PRS, we identified seven gene-trait associations with significantly earlier age-at-diagnosis in bi-allelic variants compared to heterozygous carriers of pLoF+damaging missense/protein-altering variants (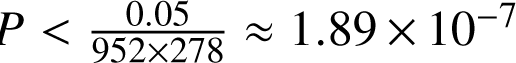, Fig. 4b-c, Supplementary Tables 16-17). Beyond these seven associations, we also identified 13 additional gene-trait relationships at a false discovery rate (FDR)< 5% (4b, Supplementary Fig. 22). For six out of the seven Bonferroni significant gene-trait combinations, we found no evidence (𝑃 > 0.05/7 ≈ 0.00833) that carrying a single heterozygous variant altered lifetime disease risk compared to carrying two copies of the reference allele.

**Fig. 4.**
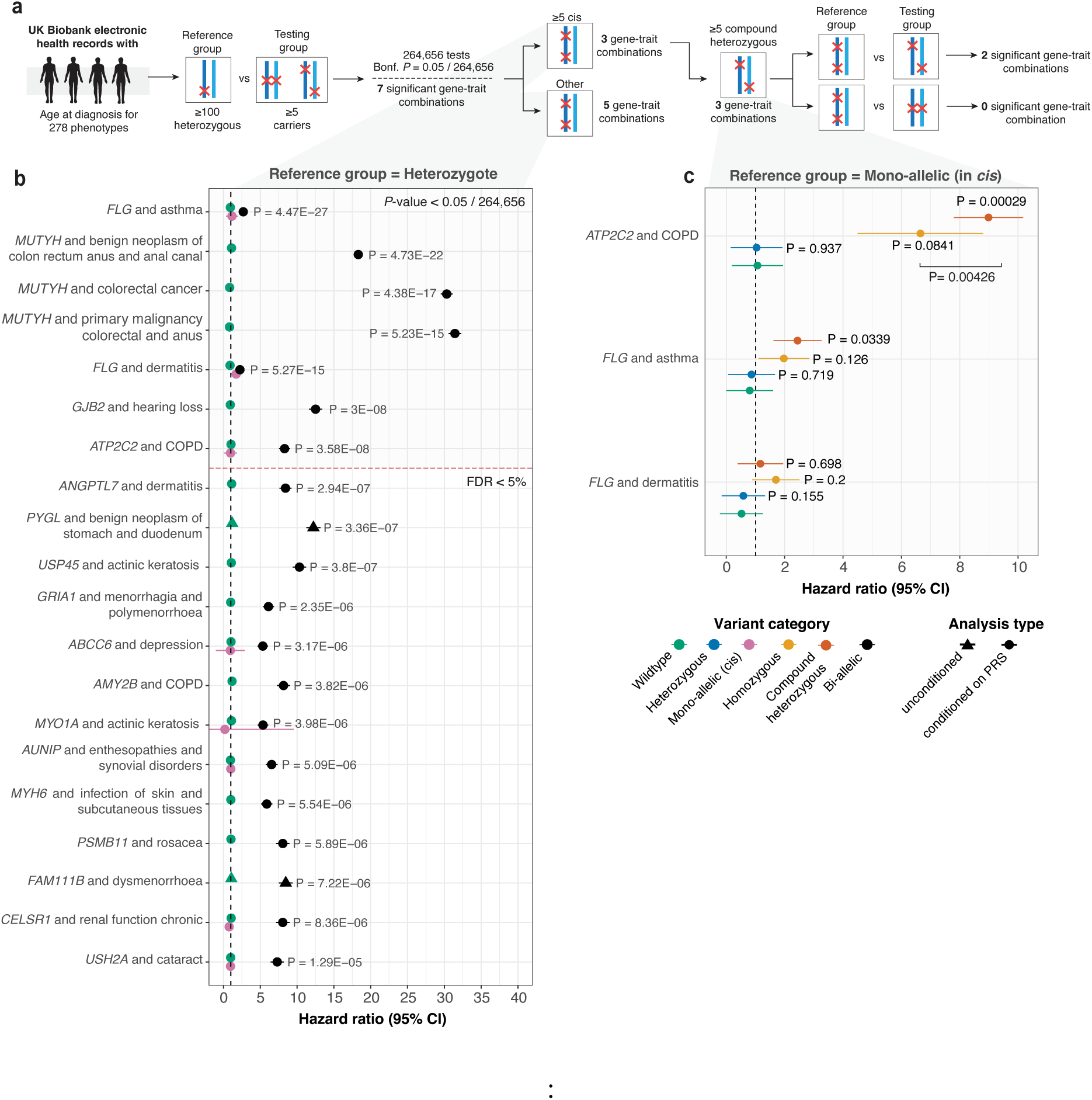
*(previous page)*: Age-at-diagnosis modeling reveals novel recessive effects driven by damaging bi-allelic variants. a) Flow diagram of our approach. To investigate whether homozygous and/or CH effects are associated with a difference in lifetime risk of disease development, we perform Cox proportional-hazards modeling for gene-trait combinations in which ≥ 5 samples are two-hit carriers (CH or homozygotes) and ≥ 100 samples that are heterozygotes. Among Bonferroni significant associations (𝑃 < 1.89 × 10^−7^), we filter to gene-trait pairs for which at least five samples carry multiple variants disrupting the same haplotype, and test for an association between CH or homozygous carrier status and lifetime disease risk (corresponding to HRs>1). b) HRs when comparing CH and homozygous status versus heterozygous carrier status. Throughout, we display hazard ratios and corresponding 𝑃-values with (circles) and without (triangles) taking the polygenic contribution into account by conditioning on off-chromosome PRSs for heritable traits that pass our quality control cutoffs. 𝑃-values following inclusion of polygenic contribution to disease status are provided where PRS are predictive. HRs for gene-traits with two or more individuals with multiple *cis* variants on the same haplotype are displayed in pink. Associations that pass Bonferroni significance (𝑃 < 1.89 × 10^−7^) and FDRs < 5% cutoff are illustrated in the top and bottom respectively. c) HRs when comparing bi-allelic status versus heterozygous carrier status for gene-trait pairs with ≥ 3 individuals harboring variants disrupting the same haplotype, allowing ascertainment of confidence intervals. c) HRs when comparing wildtype, heterozygous, CH and homozygous status against individuals that harbor two damaging variants on the same haplotype. 95% CIs are shown in the figure. Abbreviations: CC (colorectal cancer), COPD (chronic obstructive pulmonary disease).

We further sought to disentangle the effects of homozygous and CH variants on lifetime disease risk from that attributable to multiple damaging rare variant effects on a single haplotype. To do this, we analyzed such effects in the three gene-trait pairs with both (1) at least five CH and/or homozygous variants and (2) at least five individuals harboring ≥2 variants on the same haplotype (Fig. 4d, Supplementary Table 18). Compared to individuals with a single disrupted haplotype, both homozygous and CH carriers of pLoF+damaging missense/proteinaltering variants in *ATP2C2* were at increased lifetime risk of developing chronic obstructive pulmonary disease (COPD) (homozygote HR = 6.65 (95% CI = 4.5–8.8); 𝑃 = 0.084, CH HR = 8.98 (7.79–10.17); 𝑃 = 0.00028). Similarly, both homozygous and CH variants of *FLG* were at increased lifetime risk of asthma (homozygote HR = 1.97 (1.1–2.84); 𝑃 = 0.126, CH HR = 2.44 (1.61 − 3.26); 𝑃 = 0.033) and dermatitis (homozygote HR = 1.7 (0.88–2.5); 𝑃 = 0.20, CH HR = 1.16 (0.38–1.94); 𝑃 = 0.7) (Fig. 4c). For these gene-trait relationships, information encoded in genetic phase influences the risk of disease, with mono-allelic disruption leading to virtually unaltered risk while bi-allelic disruption may result in dramatic increase in lifetime risk of disease.

### Biological insights into common complex disorders implicated by CH variation

Six of the seven gene-trait combinations for which we identify Bonferroni significant associations with lifetime disease risk are also significant in our cross-sectional recessive association analysis (Supplementary Table 19). All six have previously been reported in the literature, albeit without age-at-onset effects. These include *MUTYH* and colorectal cancer, *GJB2* and hearing loss, and a pleiotropic association of *FLG* with both dermatitis and asthma. *ATP2C2*-COPD is a novel candidate association with plausible mechanistic effects.

*MUTYH*-associated polyposis is considered a highly penetrant Mendelian cancer syndrome leading to adenomatous polyposis^42^. We link bi-allelic variants of *MUTYH* to elevated risk of benign neoplasms of the colon, with bi-allelic carriers having a median age of diagnosis at age 53.7 years (interquartile range (IQR) = 47.9 - 56.3 years), as compared to heterozygotes (median age-at-diagnosis = 61.7 (56.2 - 66.7) years) and wildtypes (61.1 (54.6 - 66.5) years); as well as malignant neoplasms of the colon (median age at diagnosis for bi-allelic carriers = 52.1 (IQR = 48.6 - 53.4) years, heterozygotes = 63.2 (57.7 - 67.0) years, and wildtypes = 62.9 (57.2 - 67.9) years) (Fig. 5a-b). Because benign growths can be precursors to malignant neoplasms, and since risk of both disorders was elevated in *MUTYH* bi-allelic carriers (benign HR = 18 (95% CI = 17.72–18.9); 𝑃 = 4.7 × 10^−22^, malignant HR = 31.4 (95% CI = 30.57–32.3); 𝑃 = 5.2 × 10^−15^), we examined the co-occurrence of variants across colorectal cancer outcomes. The same set of CH and homozygous variants are involved in the pathophysiology of benign and malignant neoplasms of the colon, suggesting that *MUTYH*-variant composition alone in insufficient to explain the dichotomy between malignant and benign polyposis (Supplementary Fig. 17).

**Fig. 5.**
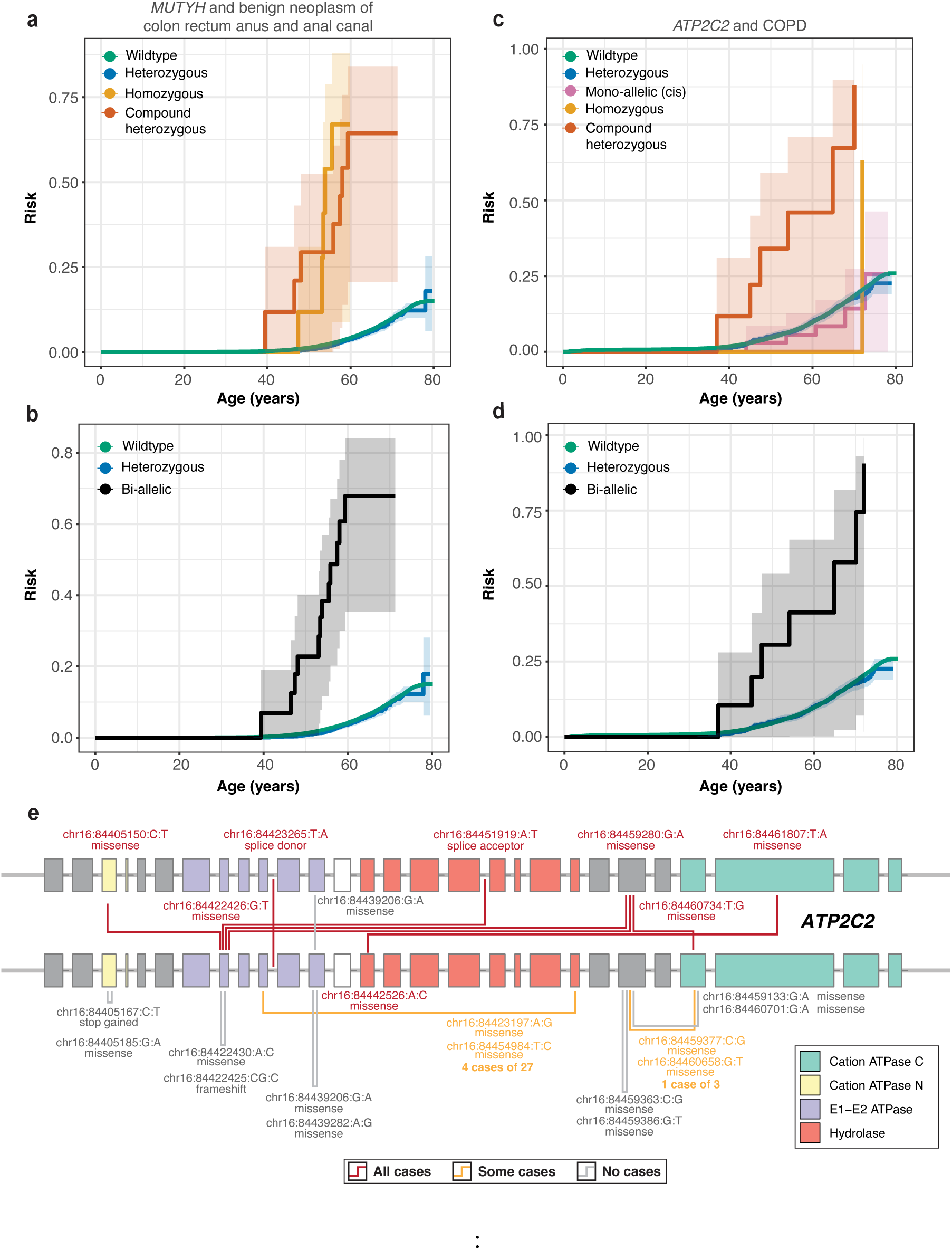
*(previous page)*: Trajectories of haplotype disruption in common disease. a-b) Kaplan-Meier survival curves for CH (red), homozygous (orange), heterozygous carriers (blue), single disruption of haplotypes (pink) owed to pLoF or damaging missense/protein-altering mutations. Wildtypes and bi-allelic variants (CH or homozygous) are shown with green and black lines respectively. Both CH and homozygous *MUTYH*-variant carriers are at elevated lifetime risk of developing benign neoplasm of the colon compared to heterozygous carriers and wildtypes. c-d) Kaplan-Meier survival curves for *ATP2C2* mono and bi-allelic variant carriers. Carriers of CH variants develop COPD more early compared to heterozygotes carriers and wildtypes. Moreover, individuals who harbor a single putatively disrupted haplotype owed to ≥2 damaging variants develop COPD at the same frequency as heterozygotes and wildtypes. e) Gene plots for *ATP2C2*, displaying protein coding variants for samples that carry ≥ 2 pLoF or damaging missense/protein-altering variants stratified by exon or intron. CH variants, multiple variants in *cis*, and homozygous variants are highlighted by lines joining the positions of co-occurring variants in a sample. Lines are colored by number of cases for the shown variant configurations, with gray lines indicating no observed samples are cases; orange lines indicating some some samples are cases; red lines indicate that all observed samples are cases. Variants are labeled by position (GRCh38) and according to inferred consequence (missense, stop gain, splice acceptor/donor). Protein domains are highlighted accordingly^44^.

ATP2C2, a calcium-transporting ATPase linked to surfactant protein D levels^43^ (a causal risk factor for COPD), is associated with COPD in our gene-trait analyses (HR = 8.3 (95% CI = 7.54–9.05); 𝑃 = 3.56×10^−8^). As we did not observe any nearby (1 Mb upstream or downstream) common variants in *ATP2C2* associated with cross-sectional COPD (all 𝑃 > 5 × 10^−6^), the association between bi-allelic variants of *ATP2C2* and COPD is potentially driven by the unique configurations of damaging-missense (𝑛 = 7) and pLoF (𝑛 = 1) variants that primarily reside in functional protein domains (Fig. 5e, Supplementary Fig. 18, Supplementary Table 20). 7 of 8 (87.5%) identified bi-allelic carriers of *ATP2C2* (6 CH and 2 homozygous) were diagnosed with COPD (median age of diagnosis = 54.1 (IQR = 46.2 - 67.5) years) (Fig. 5c-d). In contrast, only 6.9% of individuals harboring multiple pLoF+damaging missense/protein-altering variants on the same *ATP2C2* haplotype were diagnosed with COPD, and at the same median age (60.8 (53.7 - 67.9) years) as heterozygotes (58.0 (48.5 - 64.1) years) and wildtypes (59.2 (51.3 - 65.1) years).

*FLG* plays a pivotal role in the differentiation and maintenance of skin barriers^34^. *FLG* variants have been selectively associated with individuals with both asthma and atopic dermatitis, but not with those who have asthma without atopic dermatitis^35^. The exact nature of this relationship remains unclear. Our findings indicate that individuals carrying a single deleterious *FLG* allele face increased risk of dermatitis (𝑃 ≈ 7.2 × 10^−5^), but not asthma (𝑃 = 0.018), when compared to wildtypes. In contrast, individuals carrying two variant alleles have an increased risk of developing both dermatitis (𝑃 = 5.27 × 10^−15^) and asthma (𝑃 = 4.47 × 10^−27^), suggesting a recessive mode of inheritance for *FLG*-related asthma and a semi-dominant inheritance pattern for *FLG*-related dermatitis^29^. This implies that the loss of a single *FLG* copy can result in dermatitis, while the loss of both copies can lead to asthma. Together, this may help clarify why *FLG*-related asthma is seldom observed without the presence of *FLG*-related dermatitis.

## Discussion

In this large biobank-scale effort, we systematically interrogate the role of bi-allelic coding variants in genes conferring risk for common complex diseases. In the cross-sectional and longitudinal analysis we identify 20 nominally significant (𝑃 < 5.25 × 10^−5^) and 23 significant (FDR < 5%) gene-trait associations, respectively. Together, we find 36 unique gene-trait associations, that both replicate established relationships and identify previously unreported gene-trait associations for a range of binary phenotypes across the common disease spectrum.

We show that the 90% of deleterious CH variants occur at MAF < 0.34%. Given that phasing quality is directly correlated with allele frequency, it is essential to filter to the set of variants phased at high confidence to eliminate false positive identifications. Here, we quantified the increase in phasing quality using Mendelian inheritance logic in parent-offspring relationships and compared pairs of statistically phased variants to read-backed phased variants using short read sequences. While read-backed phasing is computationally expensive and restricted to variants in close proximity, we demonstrate that it can be employed to evaluate statistical phasing quality in cohorts that lack trio relationships, with error rates comparable to that of trio switch error rates.

CH disease associations have mainly been explored in rare disorders^13–17^, but are seldom investigated in the study of common disease. This is due to the low prevalence of variants in the CH state and the genetic architecture of common complex traits, which are typically influenced by environmental factors and numerous loci with low to modest contribution to risk. In this study, we address these challenges and offer multiple lines of evidence to demonstrate the role of CH effects in driving disease risk for common traits. We employed two complementary analyses to detect gene-trait associations: a genome-wide logistic association analysis and a time-to-event model. Through these methods, we identified associations in which variants in the homozygous or CH state resulted in increased disease risk compared to wildtypes and individuals carrying multiple pathogenic variants on the same haplotype. Our findings show that for certain gene-trait pairs, individuals with a single disrupted gene copy have a risk of developing disease that is virtually indistinguishable from that of wildtypes, suggesting non-additive gene dosage effects. Further, by permuting the genetic phase, we find evidence that incorporation of confidently phased CH variants can boost power to detect recessive associations in common disease. Collectively, our results emphasize the importance of considering each individual’s specific genetic context when assessing their genetic risk in a clinical setting. Simply identifying the presence of multiple pathogenic variants in a gene, disregarding the phase, may not be sufficient to fully understand an individual’s risk profile.

Many common complex traits have polygenic architectures, which should be accounted for when performing gene-trait association testing. The presence of bi-allelic variants in individuals with such diseases might be coincidental and not causally related to the trait, which may instead be a result of a high polygenic risk. However, across the significant recessive genome-wide associations, we observed that inclusion of PRS as a covariate, affected the resulting association *P*-value by less than single order of magnitude for the binary traits we analyzed. While we were only able to account for the polygenic contribution to disease development for 111 diseases with significant common variant heritability in the UKBB, due to low case numbers, these observations suggest that incorporation of polygenic background has limited influence on the degree of association when evaluating ultra-rare variation across binary traits.

We found that the majority of bi-allelic gene-disease associations are driven by variant combinations containing at least one missense variant, which would have been excluded under a stricter high-confidence pLoF criterion. Although our less stringent inclusion threshold enabled us to identify a greater number of bi-allelic variants, it is likely that some damaging missense or protein-altering variants would incorrectly be predicted as damaging, or may exhibit gain-of-function rather than loss-of-function effects, consequently reducing the signal-to-noise ratio in our analyses. Even ‘knockouts’ by *bona fide* pLoF variants may only result in partial gene inactivation, and not necessarily complete gene knockdown. Additionally, pLoF variants may be ‘rescued’ and not lead to complete or even partial loss-of-function. While we show that including damaging missense/protein-altering variants to define bi-allelic variants can improve power for certain phenotypic associations, further manual curation and experimental validation will be required to demonstrate that these variants truly result in loss-of-function.

The likelihood of damaging alleles occurring on the same haplotype is influenced by a complex interplay of factors, including population structure and balance between selection, drift, mutation, and recombination. We and others^45^ find that damaging CH variants occur less frequently than multiple damaging variants affecting the same haplotype, suggesting that in certain circumstances, natural selection operates on a haplotype level. Once a LoF variant occurs and expands in the population, the affected haplotype has no selection against additional acquisition of damaging mutations. This has implications for association studies investigating CH effects by counting the number of damaging variants in a gene while attributing equal probability to each of affecting each haplotype^8^, as such frameworks may overestimate the frequency of CH events.

‘Human knockouts’ have been extensively discussed in the context of therapeutic development. Examining both bi-allelic and mono-allelic carriers can help assess the safety of therapeutic interventions by analyzing how varying degrees of target modulation affect biological response^3, 6^. We showcase several gene-trait relationships where the number of affected haplotypes influences the lifetime risk of disease, potentially representing the manifestation of ‘adverse events’ which are important endpoint in clinical trials. The absence of adverse events in mono-allelic carriers can potentially imply that partial pharmacological inhibition of a target may be a safe and effective approach. However, adverse effects observed in bi-allelic carriers of damaging variation within the same locus could indicate potential risks associated with complete target inhibition. A natural extension of this work could involve investigating mono and bi-allelic effects on quantitative outcomes, such as serum proteins. Changes in biomarkers (or other continuous outcomes) may reflect direct or indirect consequences of gene modulation and could serve as potential pharmacodynamic biomarkers commonly used to assess target engagement in clinical trials.

This work showcases the value of statistical phasing of damaging rare variants, and that association analyses that account for compound heterozygosity can be better-powered for gene-trait discovery. We show that this approach can be employed to discover well-established and novel non-additive and additive gene-trait relationships across a wide range of disease etiologies. From a clinical perspective, we demonstrate the importance of interrogating the genetic phase when dealing with CH variants in traits with recessive mode of inheritance. This is an important step towards uncovering the phenome-wide consequences of bi-allelic disruption across the human genome.

## Code availability

The code required to reproduce our analyses are publicly available at https://github.com/ frhl/wes_ko_ukbb. Data produced in the present study are available upon reasonable request to the authors.

## Data Availability

The code required to reproduce our analyses is publicly available at https://github.com/frhl/wes_ko_ukbb. Data produced in the present study are available upon reasonable request to the authors.

## Acknowledgments

F.H.L. is supported by the Wellcome Trust (award 224894/Z/21/Z), and the Medical Sciences Doctoral Training Centre at the University of Oxford. S.S.V. is supported by the Rhodes Scholarship, Clarendon Fund, and the Medical Sciences Doctoral Training Centre at the University of Oxford. N.B. is supported by the Clarendon Fund, and the Medical Sciences Doctoral Training Centre at the University of Oxford. W.Z. is supported by the National Human Genome Research Institute of the National Institutes of Health under award number K99HG012222. A.B. is supported by the Novo Nordisk Center for Genomic Mechanisms of Disease at the Broad Institute (NNF21SA0072102). N.W. is supported by a Sir Henry Dale Fellowship jointly funded by the Wellcome Trust and the Royal Society (220134/Z/20/Z) and research grant funding from the Rosetrees Trust (PGL19-2/10025). C.M.L. is supported by the Li Ka Shing Foundation, NIHR Oxford Biomedical Research Centre, Oxford, NIH (1P50HD104224-01), Gates Foundation (INV-024200), and a Wellcome Trust Investigator Award (221782/Z/20/Z). This research has been conducted using the UK Biobank resource under application Number 10844.

## Methods

### Exome sequencing quality control summary

We perform a series of hard-filters on genotype, sample, and variant metrics (Table 1, Table 2-3). We confirm genetic sex with reported sex, and restrict analysis to genetically ascertained samples of NFE ancestry, using random forest (RF) classifiers (Fig. 2-3). Finally, we filter based on a second collection of sample and variant filters (Tables 2-3). We used Hail 0.2^46^ and PLINK 1.9^47^ to perform all QC steps, and use R (4.0.2) scripts for plotting and filtering. Data was manipulated in R using data.table (1.14.2) and dplyr (1.0.7), random forest classifiers were trained using the randomForest (4.6-14) library, and plotting was performed using a ggplot2 (3.3.5).

### Exome sequencing quality control, full details

#### Sample filters

We evaluated sample-level quality control (QC) metrics on the 200,643 UKBB ES multi-sample project level variant call format (VCF) call-set files^46^, Supplementary Table 1. All metrics were calculated for bi-allelic single nucleotide polymorphisms (SNPs), except for metrics involving insertions and deletions. We regressed out the first 21 principal components (PCs)^48^, and filtered out sample outliers of the residuals for each metric based on MAD (median absolute deviation) thresholds (Supplementary Table 1). Samples without PC data were subject to more stringent thresholds (Supplementary Table 1).

**Supplementary Table 1:** Sample filtering: MAD Intervals. The interval [𝑎, 𝑏] represents median(𝑋) + MAD(𝑋) [−𝑎, 𝑏] for the metric, 𝑋. Samples with metrics outside these intervals were removed.

| **Metric** | **Metric residual (w/ PCs)** | **Raw (w/o PCs)** |
| --- | --- | --- |
| *call_rate* | [24, ∞) | [4, ∞) |
| *n_insertion* | [8, 8] | [4, 4] |
| *n_deletion* | [8, 8] | [4, 4] |
| *r_insertion_deletion* | [8, 8] | [4, 4] |
| *n_het* | [12, 12] | [4, 4] |
| *n_hom_var* | [12, 12] | [4, 4] |
| *r_het_hom_var* | [16, 16] | [4, 4] |
| *n_non_ref* | [8, 8] | [4, 4] |
| *n_singleton* | $(-\infty, 16]$ | $(-\infty, 4]$ |
| *n_snp* | [8, ∞) | [4, 4] |
| *n_transition* | [8, 8] | [4, 4] |
| *n_transversion* | [8, 8] | [4, 4] |
| *r_ti_tv* | [8, 8] | [4, 4] |

#### Genotype filters

Multi-allelic variants were split into bi-allelic variants and insertions and deletions (indel) were left-aligned^49^. Genotype calls meeting any of the following criteria were set to missing:

1. Genotype quality (GQ) ≤ 20
2. Total sequencing depth (DP) ≤ 10
3. Heterozygous calls:
  a. SNPs: 1-sided binomial test of alternate allele depth related to total read depth 𝑃 < 1 × 10^−3^
  b. Indels: alternate allele read depth / total read depth < 0.3
4. Homozygous indel calls: alternate allele read depth / total read depth < 0.7

#### Variant-level filters

Retain variants satisfying all of the following conditions:

1. Not in a low complexity region (LCR)^50^.
2. In sequencing target regions ±50 base pairs.
3. MAF > 0 following genotype QC.
4. Excess heterozygosity (ExcessHet < 54.69) filter: Phred-scaled 𝑃-value for exact test of excess heterozygosity^51^ in founders as determined by relatedness estimates and recorded ages of UKBB participants^48^. Variants were retained as recommended in genome analysis toolkit (GATK)^51^

### Additional ES quality control

To perform further QC we use Hail, an open-source Python library which focuses on the analysis of large-scale genetic data sets. We used Hail to create our own methods, and we take advantage of the functionality that has been rewritten to enable fast and scalable analysis of large exome and genome sequencing projects. Unless otherwise stated, all of the following the data curation and quality control steps were performed in Hail^46^.

Briefly, we apply a collection of hard-filters on sample metrics. We confirm genotypic sex with reported sex, remove samples with excess glsplurv, and restrict analysis to samples of genetically ascertained NFE ancestry. Finally we apply a second collection of sample and variant hard filters. As an initial pass to remove low quality and contaminated samples, we filter out samples with call rate < 0.95, mean DP < 19.5× or mean GQ < 47.8 (Fig. 1).

### Sex imputation

To confirm participant sex and calculate PCs, we extracted high quality common variants (allele frequency between 0.01 to 0.99 with high call rate (> 0.98)) and LD prune to pseudo-independent SNPs using --indep 50 5 2 in PLINK 1.9. When reported sex does not match genotypic sex, it may signal potential sample swaps in the data. Using the 𝐹-statistic for each sample using the subset of the non-pseudo autosomal region on chromosome X, we identify and remove samples where reported sex information is not confirmed in the sequence data (Fig. 2). Specifically, we remove samples satisfying at least one of the following criteria:

- Sex is unknown in the phenotype files.
- 𝐹-statistic > 0.6 and the sex is female in the phenotype file.
- 𝐹-statistic < 0.6 and the sex is male in the phenotype file.
- 𝐹-statistic > 0.6 and number of calls on the Y chromosome is < 100.

**Supplementary Fig. 1:**
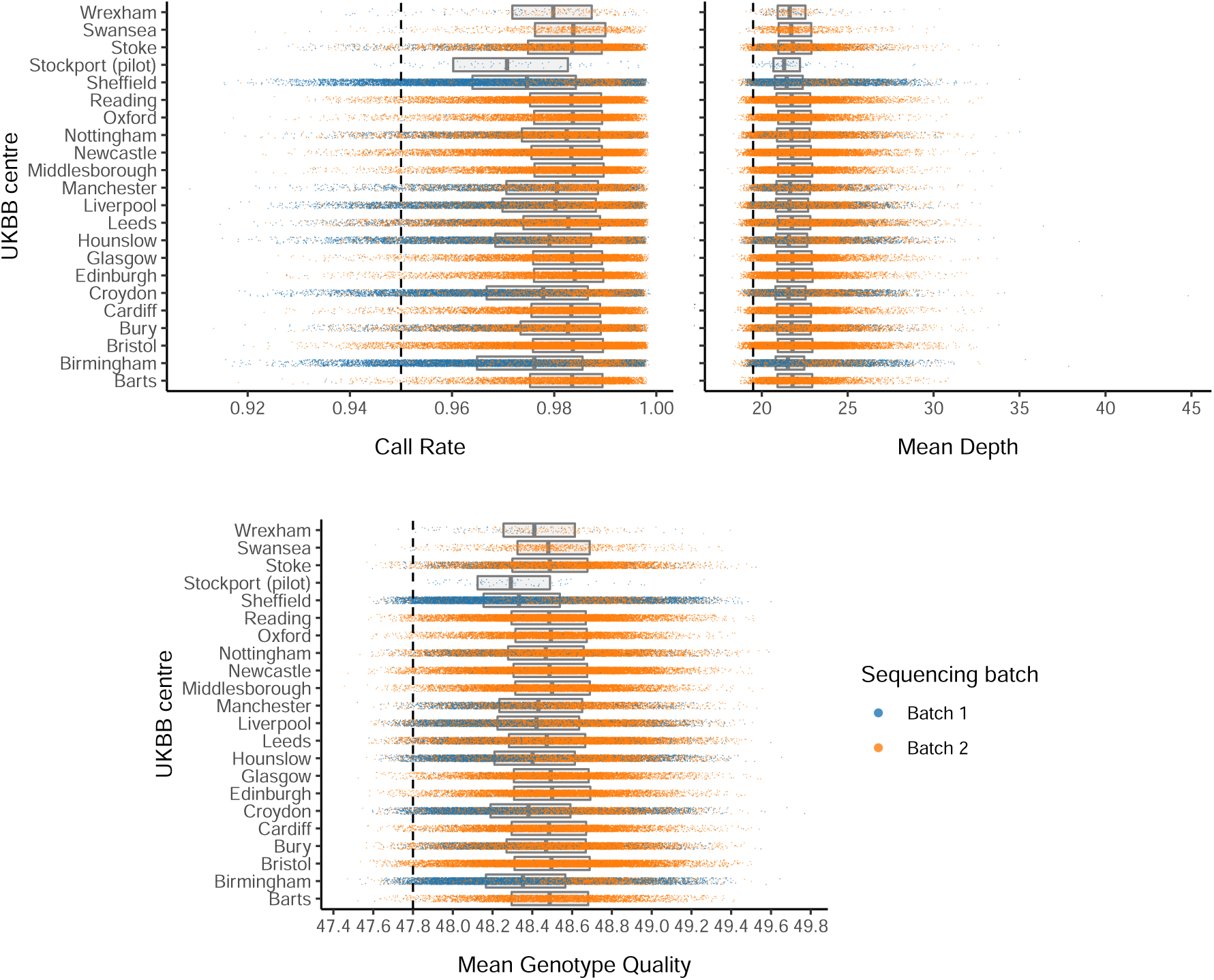
Distributions of sample metrics following initial restriction to variants, lying outside LCRs and inside the padded (50 bp) target intervals, and prior to the initial hard sample filters (call rate > 0.95, mean depth > 19.5, mean GQ > 47.8). In each plot, jittered scatters display the distribution for each UKBB recruitment center, colored according to sequencing batch. Box-plots behind the scatter display the median and interquartile range for each sequencing batch. Hard-filtering thresholds are denoted by the dashed vertical line.

**Supplementary Table 2:**
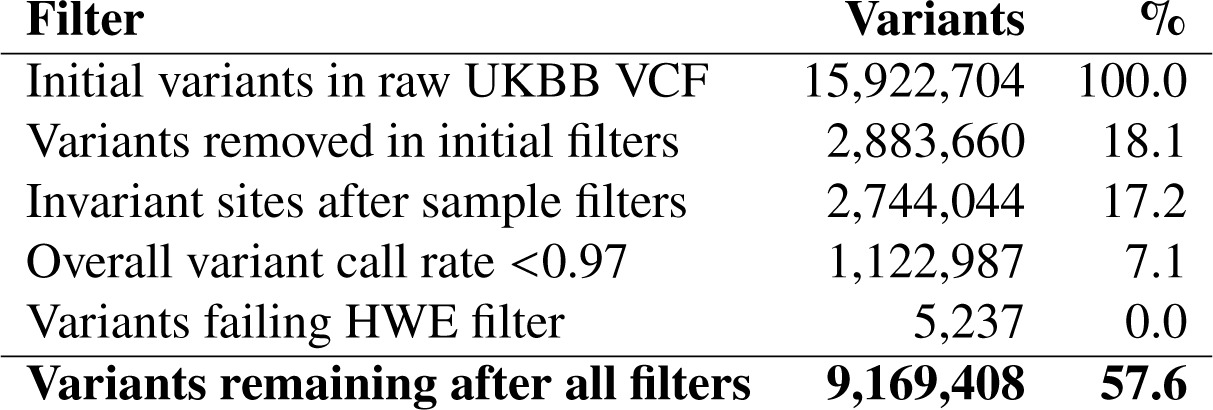
Summary of sample filters. Moving down through the rows of the table, we move through QC filtering steps.

**Supplementary Table 3:**
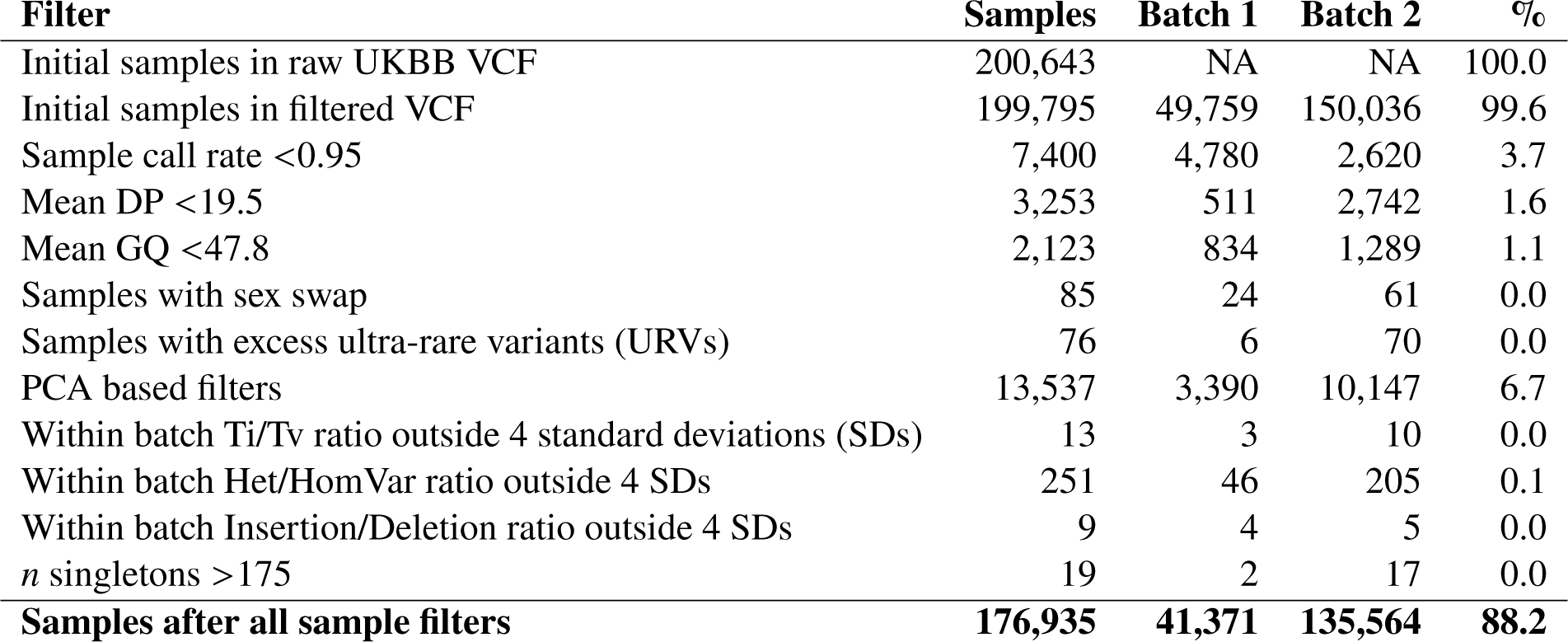
Summary of variant filters. Moving down through the rows of the table, we move through QC filtering steps.

### Defining a set of samples with non-Finnish European ancestry

To ensure adequate case-control for as many traits as possible, we restricted our analysis to a set of genetically ascertained NFE samples. To do this, we perform a number of principal component analysis (PCA) steps to ensure that we have subset down to NFE. We first run PCA on the 1000 Genomes (1KGP) samples (minus the small subset of related individuals within the 1KGP) using subsetting to LD pruned autosomal variants. We then project in the UKBB samples, ensuring that we correctly account for shrinkage bias in the projection^52^. Next, we removed samples outside of the European population (EUR) using a RF classifier: we train a RF on the super-populations labels of 1KGP and predict the super-population for each of the UKBB samples (Fig. 3). We denote strictly defined European subset as those with probability > 0.99 of being European according to the classifier. Another RF classifier is trained following restriction of the 1KGP samples to Europeans to determine NFE, using a classifier probability of 0.95. RF classifiers were trained using the randomForest (4.6) library in R. Samples not assigned to the NFE cluster were removed from downstream analysis.

**Supplementary Fig. 2:**
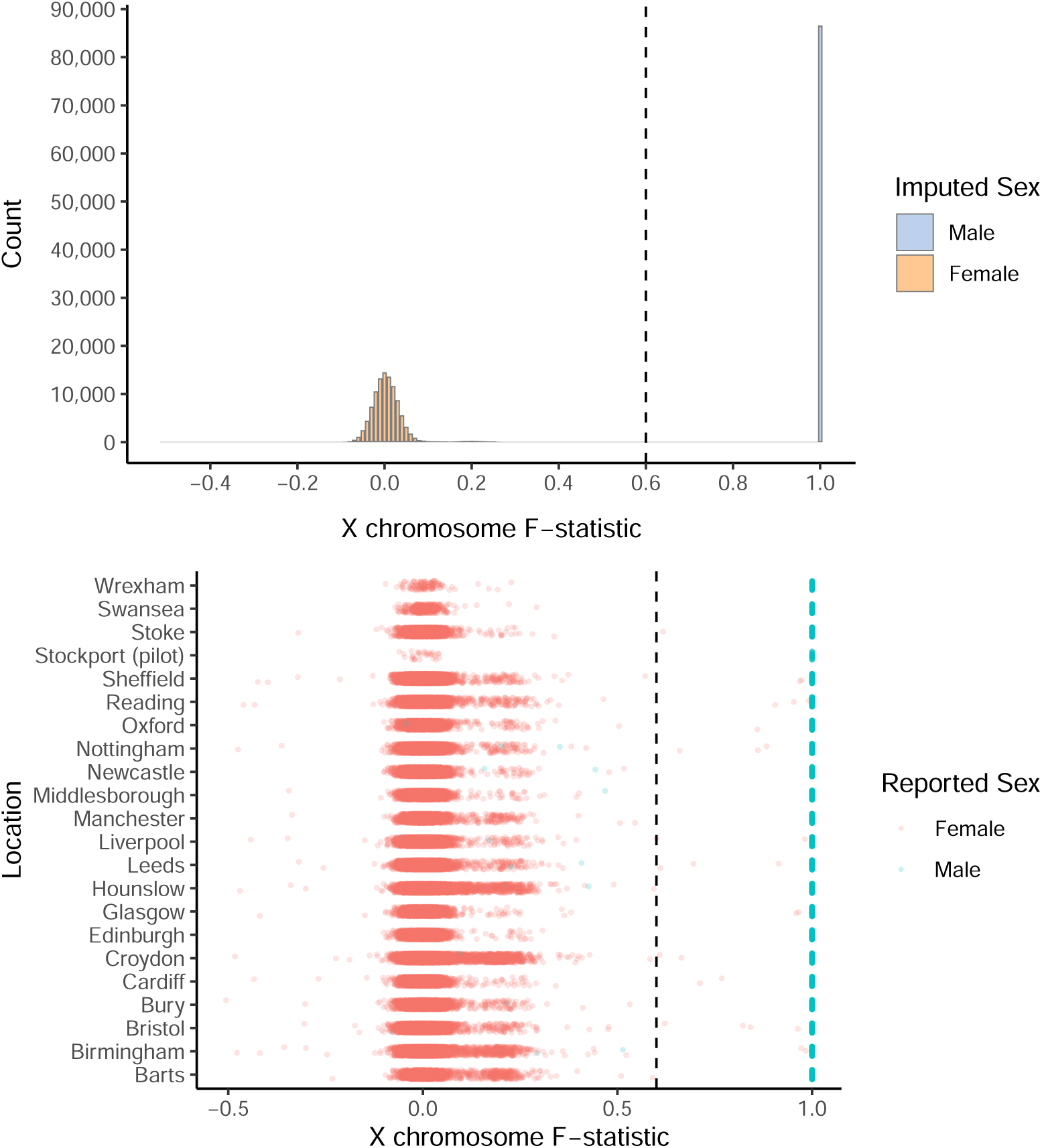
Histogram and scatter-plots of X chromosome 𝐹-statistic by collection. Samples lying to the left and right of the dashed line were called as female and male respectively, according to the imputed sex colorings in the upper histogram. Reported sex, split by UKBB recruitment center are shown in the lower jittered scatter-plots: red if the sample is reported as female, and blue if the sample is reported as male.

**Supplementary Fig. 3:**
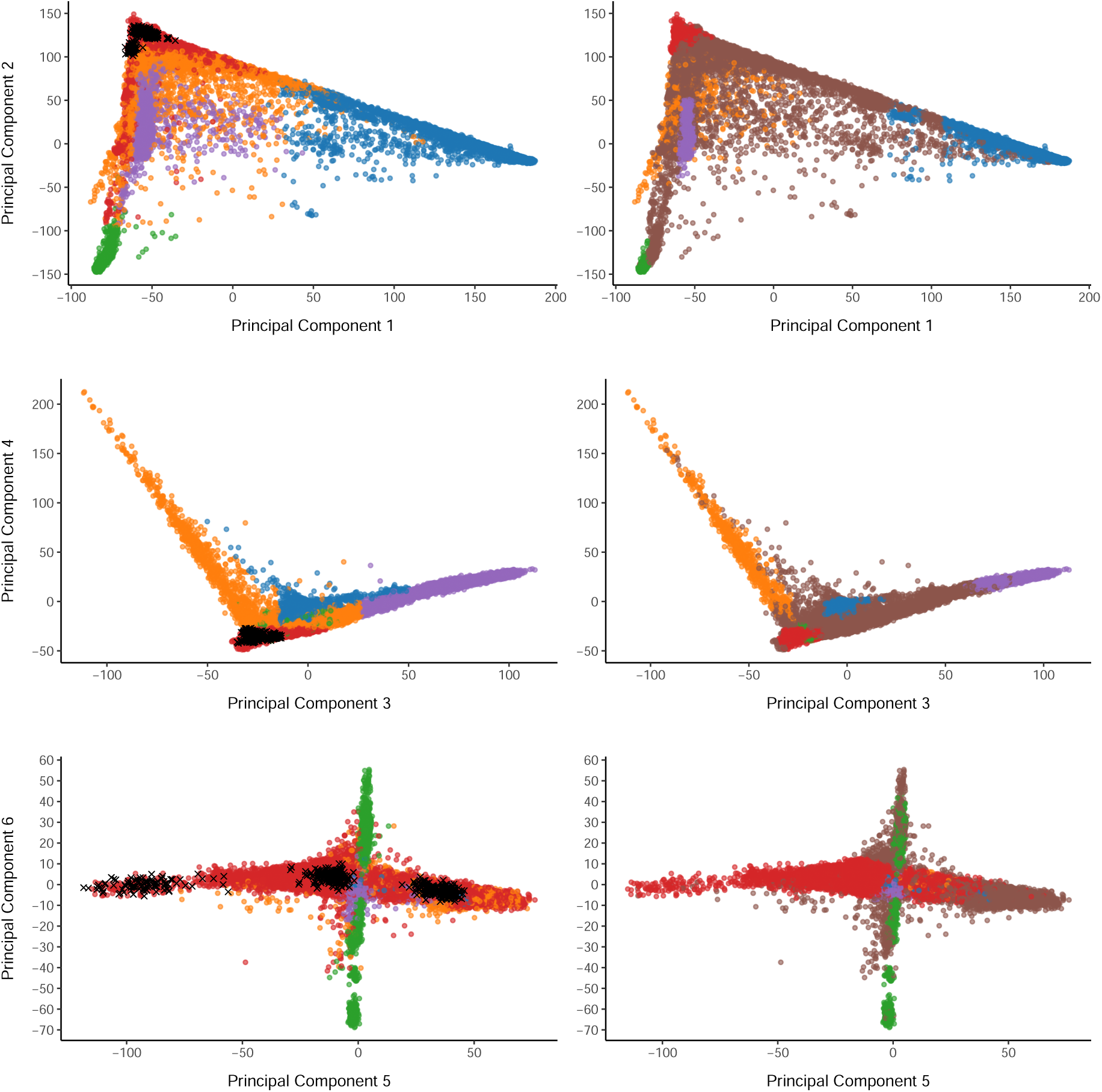
Scatter-plots of PCs of UKBB genotype data projected into the PC space defined by 1KGP samples. Points are colored according to sample collection, with 1KGP samples colored in blue. 1KGP super-populations labels were used to train a random forest classifier.

### Final hard filters

For our final variant filtering step, following restriction to the NFE subset, and removal of incorrectly defined sex or unknown sex, and run variant QC. We then filter out variants with call rate < 0.97, variants out of Hardy-Weinberg equilibrium (HWE) (𝑃 < 1 × 10^−6^), and remove invariant sites following the previous sample based filters. After restricting to these high quality variants, we perform a final set of sample filters to finalize the quality controlled data. We evaluate a collection of sample metrics and remove samples falling outside four SDs of the sequencing batch mean (Ti/Tv, Het/HomVar, Insertion/Deletion ratios), and remove the collection of samples with over 175 singletons. The resultant curated analysis ready data set consists of 176,935 samples, and 9,169,408 variants (Supplementary Table 2-3).

A summary of sample and variant filters are provided in Supplementary Tables 2-3. The high quality ES call-set consisted of 176,935 samples and 9,169,408 variants.

## Phasing

### Combining ES data with genotype array data

We combined genotyping array (UK BiLEVE Axiom array and UKBB Axiom array) and exome chip (IDT xGen Exome Research Panel v1.0) variants after general ES quality control using Hail^46^ and BCFtools^53^ (1.12). For variants in both data sets, we preferentially retained those on the ES data. For variants on the genotyping array we excluded variants missingness > 5% after performing a liftover to GRCh38 using Hail^46^. To avoid biasing the phasing quality estimates, we excluded parents among trio relationships prior to phasing. We first created a common variant scaffold by phasing variants in the combined (exome sequencing and genotyping array) data with MAF > 0.1% and otherwise default parameters using SHAPEIT5 PHASE COMMON module. We then phased the remaining rare variants using the common variant scaffold using the SHAPEIT5 PHASE RARE with recommended parameters. To ensure computational tractability, we phased overlapping chunks of 100,000 variants with ≥ 50, 000 variant overlap between consecutive chunks using Hail^46^. Following chunk phasing, we then removed the initial and final 22,500 variants from each chunk, so that 5,000 overlapping variants remained between contiguous phased chunks. We then combined the phased chunks, matching haplotype phase using bcftools^53^ (1.12) with the --ligate option. We then restrict this phased genetic dataset to the set of samples and variants present in the analysis ready NFE subset (Supplementary Tables 2-3).

**Supplementary Fig. 4:**
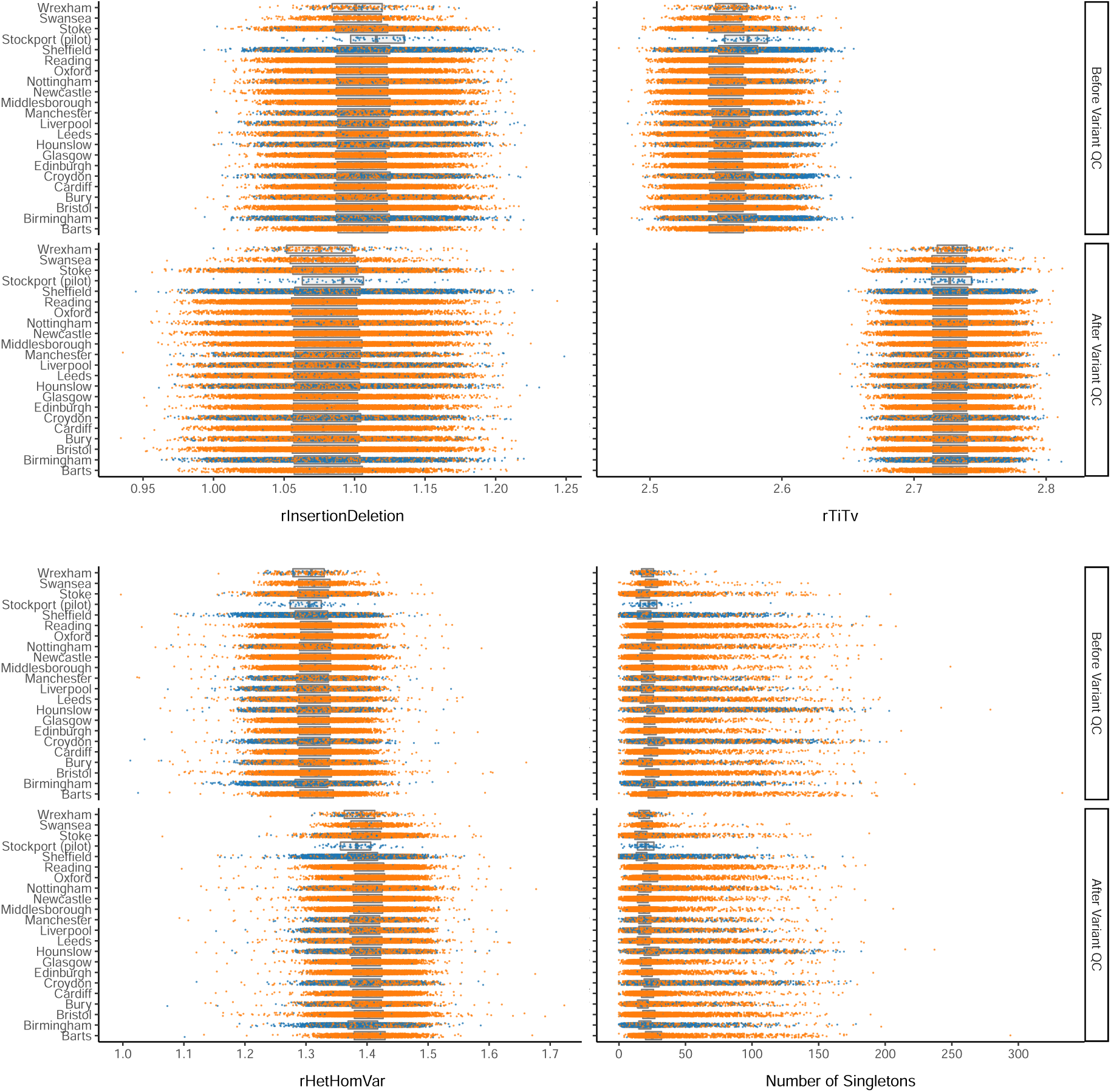
Distributions of variant metrics before and after the removal of invariant sites, variants with call rate < 0.97, and variants out of HWE (𝑃 < 1 × 10^−6^). In each plot, jittered scatters display the distribution for each sequencing batch colored by sequencing batch. Box-plots behind the scatter display the median and interquartile range for each sequencing batch. Points shown are following variants hard-filters and prior to removal of variants with metrics outside four standard deviations of the sequencing batch mean.

### Trio-switch error rates

We assessed phasing quality by comparing statistically phased genotypes to those implied in 96 trios using Mendelian inheritance logic. Switch errors are determined by traversing the statistically phased and parent-offspring transmitted haplotypes simultaneously and scanning for inconsistencies in phase between pairs of contiguous variants. This method only allows us to consider sites in which the one parent is heterozygous and the other is homozygous for the reference or alternate allele, and thus do not consider *de novo* variants or Mendelian inconsistencies in the trio data. To assess switch error in a site-specific manner, we modified and recompiled bcftools^53^ (1.12) to output errors by genomic position. We then used the modified version to assess switch by variant categories, for example by genetic data modality (genotyping array or ES), or by MAF bins. To evaluate switch errors across different phasing confidence thresholds, we filtered VCF using Hail^46^ and then repeated the switch error calculation step. We calculated binomial 95% confidence intervals (CIs) for SERs using the R-package HMisc^54^ (4.7).

**Supplementary Fig. 5:**
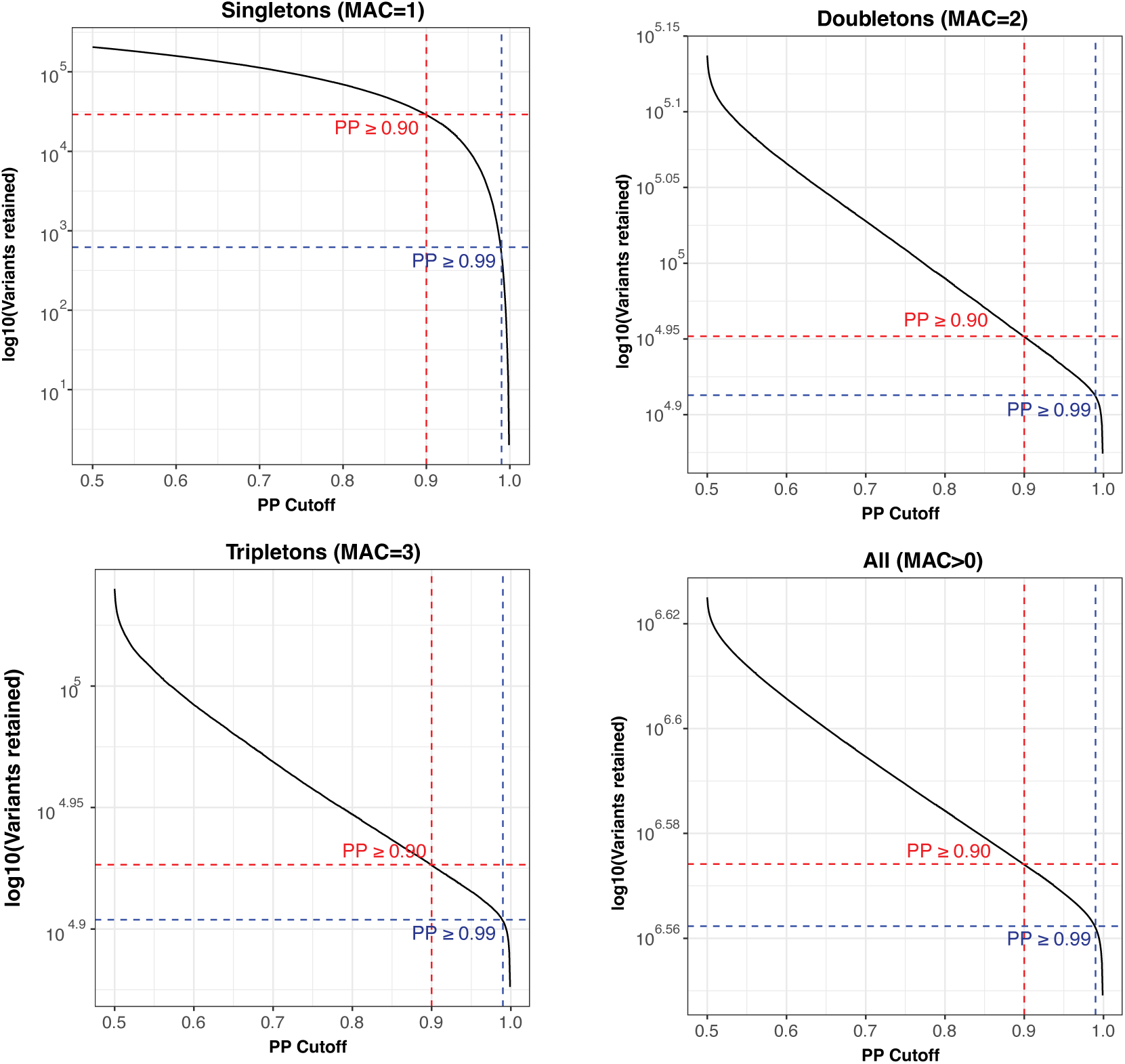
Phased variants retained as a function of phasing confidence score. Each subplot displays the number of variants retained on the log_10_ scale as the PP is increased, split by rarity of variants described in the subplot title. Dotted red and blue lines highlight the number of variants retained after imposing PP cut-offs of 0.9 and 0.99, respectively.

**Supplementary Fig. 6:**
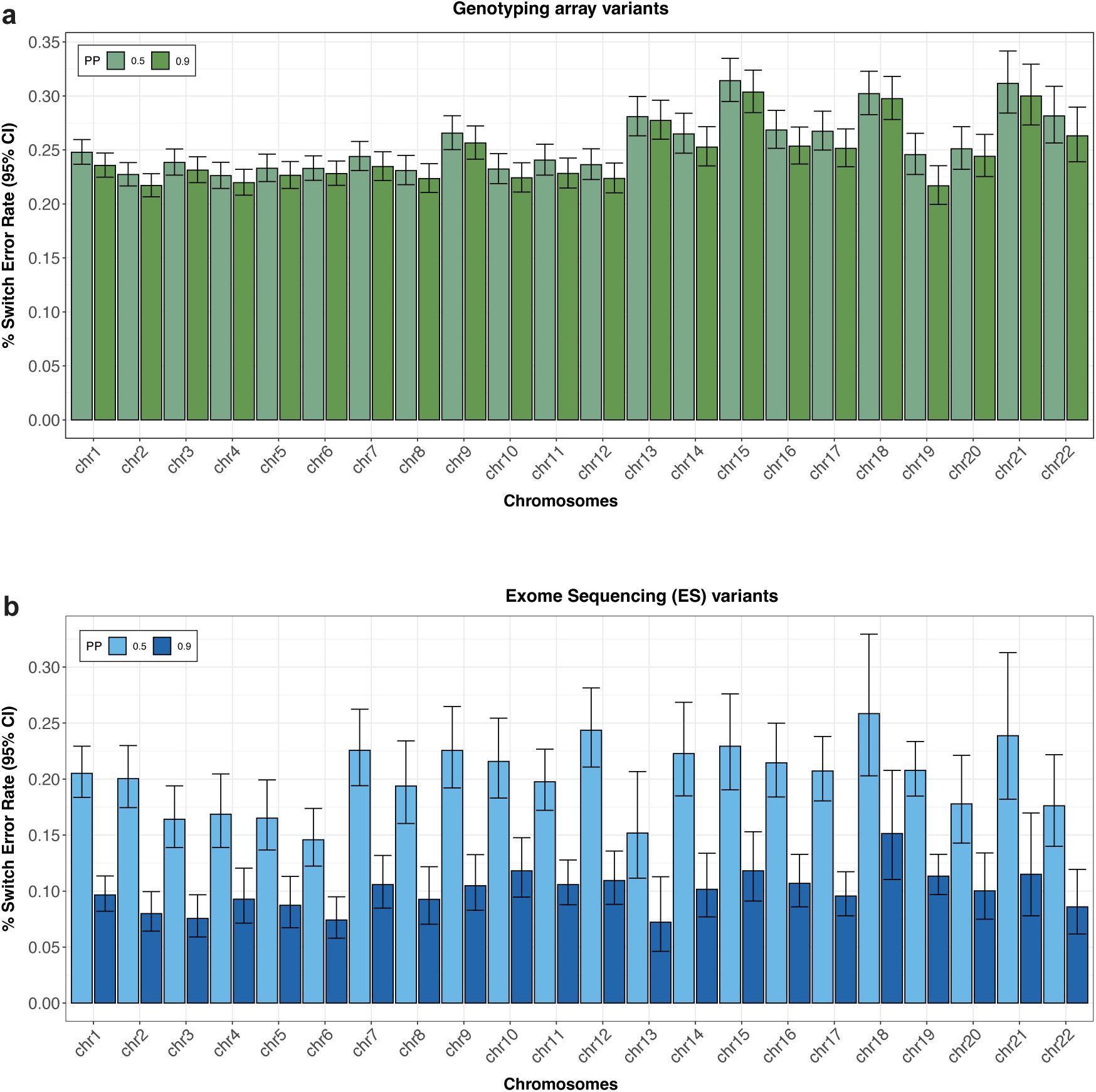
Trio switch error rates by chromosome. Parent-offspring trios are used to determine switch error rates for variants that originate from the genotyping array (a) and exome sequencing data (b). We stratify by phasing confidence (PP) according to the color legends. Mean switch error rates are plotting, with whiskers enclosing the 95% binomial CI.

### Read-backed phasing

We performed read-backed phasing with UKBB ES short paired-end read sequences using .cram files provided by UKBB. As WhatsHap is computationally expensive, we restricted our analysis to pairs of variants on chromosomes 20-22 in 176,586 genetically ascertained NFEs. We phased both single nucleotide polymorphism (SNV) and indel with WhatsHap^26^ using the default recommended parameters. WhatsHap outputs lists of phased variants within ‘phased sets’. We carried forward reads overlapping no more than two variants, for which phase could be inferred. We combined these phased variants with statistically phased variants from SHAPEIT5 using Hail^46^, and determined agreement between estimated phasing in WhatsHap and SHAPEIT5 (Fig. 7).

### Phenotype curation

We considered a collection of 282 binary quality controlled and publicly available common complex phenotypes for analysis^55^. To complement these, we also considered 28 common complex phenotypes that were obtained through manual curation, resulting in a total of 311 binary phenotypes for analysis. To increase our power for analyses for binary traits, we amalgamated a collection of phenotypes where possible: combining the phenotype curation of Censin et al.^56^, with the primary care mappings file provided by UK Biobank all lkps map v3.xlsx and our own manual curation. We aggregated across ICD-10, ICD-9, operating codes, nurses interview reports, and self-reported diagnosis by doctor from the main phenotype file, as well as v2 and v3 read codes in the primary care data. As in Censin et al., we made use of the careful definitions of Eastwood *et al.*^57^, subsequently applied by Udler *et al.*^58^ for diabetes subtype curation. Briefly, the algorithm developed in Eastwood *et al.* bins individuals into putative diabetes status using a collection of phenotypes in the UK Biobank data including self-reported diabetes diagnosis, age of diagnosis, medications, start of insulin within a year of diagnosis. We defined cases as those placed in the probable and possible case categories in the algorithms output. Controls were defined as samples labeled as ‘diabetes unlikely’ by the algorithm.

**Supplementary Fig. 7:**
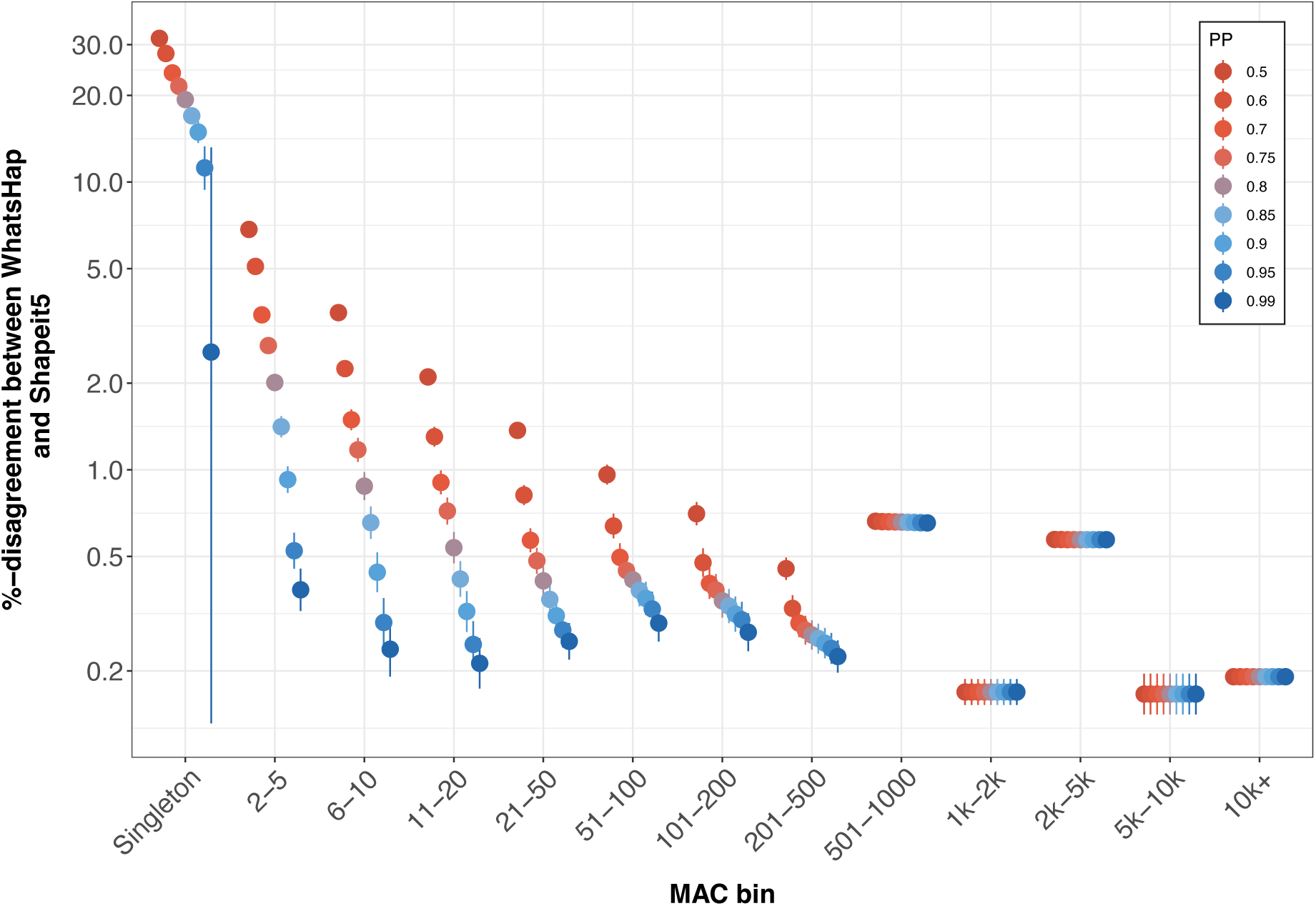
Agreement between read-backed and statistical phase estimation. Genetic phase was estimated using WhatsHap (read-backed phasing) and SHAPEIT5 (statistical phasing) in 176,586 individuals on chromosomes 20-22. We only carried forward pairs of variants close proximity in which phase could be inferred using WhatsHap. We combined with statistically phased counterparts derived from SHAPIET5 and determine % disagreement of phase estimation of variant pairs on the 𝑦-axis, when filtering to phased pairs of variants where the minimum PP > 𝑝 for 𝑝 ∈ {0.5, 0.6, 0.7, 0.75, 0.8, 0.85, 0.9, 0.95, 0.99} according to the color legend. We stratify pairs of variants into bins based on the minimum MAC in the variant pair, on the 𝑥-axis. Mean disagreement rates are plotted on 𝑦-axis with whiskers enclosing the 95% binomial CI

**Supplementary Fig. 8:**
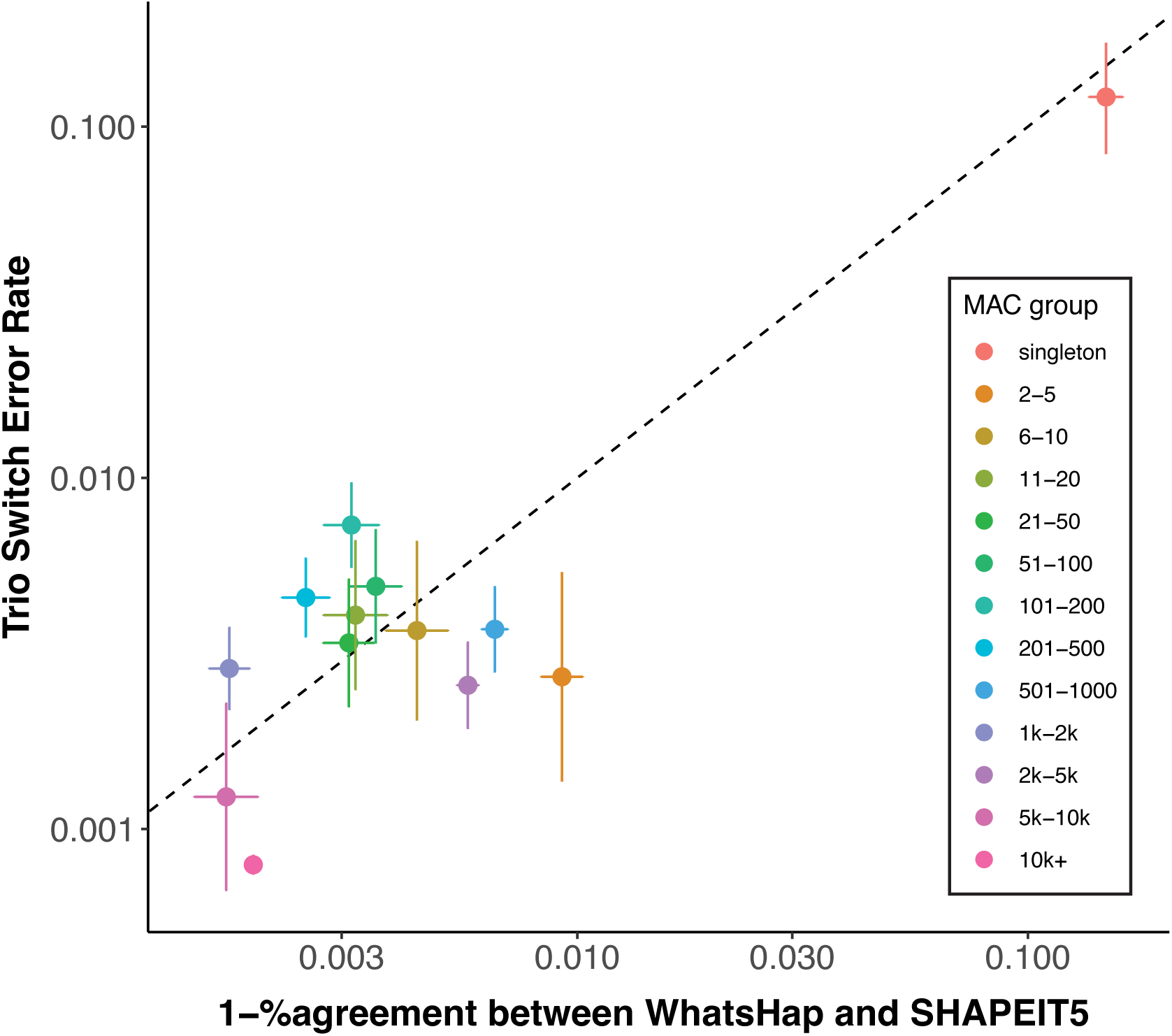
Agreement between read-backed phasing and statistical phasing. We plot the disagreement between WhatsHap (read-backed phasing) and SHAPEIT5 (statistical phasing) in UKBB on the 𝑥-axis against switch error rate in SHAPEIT5 phase estimates implied by trio-based phasing in UKBB on the 𝑦-axis. For each comparison, bin pairs of variants according to the minimum MAC in the variant pair according to the color legend. Horizontal and vertical lines enclose 95% binomial CIs around mean estimates. The dotted line is included to display 𝑦 = 𝑥.

### Variant annotation masks

We annotated coding variation using Variant Effect Predictor (VEP)^59^ (v95) using the worst consequence by gene within ‘canonical’ transcripts. We classified variants into four categories: protein truncating variants (PTVs), missense variants, synonymous variants, and other variants (Supplementary Table 9). We then split PTVs into putative loss of function (pLoF) (HC) and LC loss-of-function variants using LOFTEE^60^, and labeled missense variants with both Rare Exome Variant Ensemble Learner (REVEL)^61^ score ≥ 0.6 and CADD^62^ score ≥ 20 as ‘damaging missense’ or otherwise as ‘other missense’. Finally, we combine the resultant ‘damaging missense’ category with LC loss-of-function variants, which we denote as ‘damaging missense/protein-altering’.

### Bi-allelic encoding and recessive association modeling

Using custom Hail scripts, we define and annotate individuals as being ‘bi-allelic’ for a gene if they harbor at least one pLoFs or damaging missense variant with MAF < 5% on both inherited copies of the gene. For each sample, we encoded the presence and absence of a damaging bi-allelic variant for each gene as zero and two, respectively. We encode this information in a .vcf file and test for an association between presence of a damaging bi-allelic variant in a gene and a trait using SAIGE^63^, adjusting for sex, age, sex × age, age^2^, UKBB centre, genotyping batch and the first 10 PCs. We took relatedness into account using a sparse genetic relatedness matrix (GRM) fitted on NFE. We restrict analysis to (gene, trait) pairs with at least five bi-allelic variants in the curated ES with non-missing corresponding phenotype data (corresponding to a minimum MAC ≥ 10), and adjust for multiple testing at Bonferroni significance (𝑃 < 0.05/gene-trait pairs).

**Supplementary Fig. 9:**
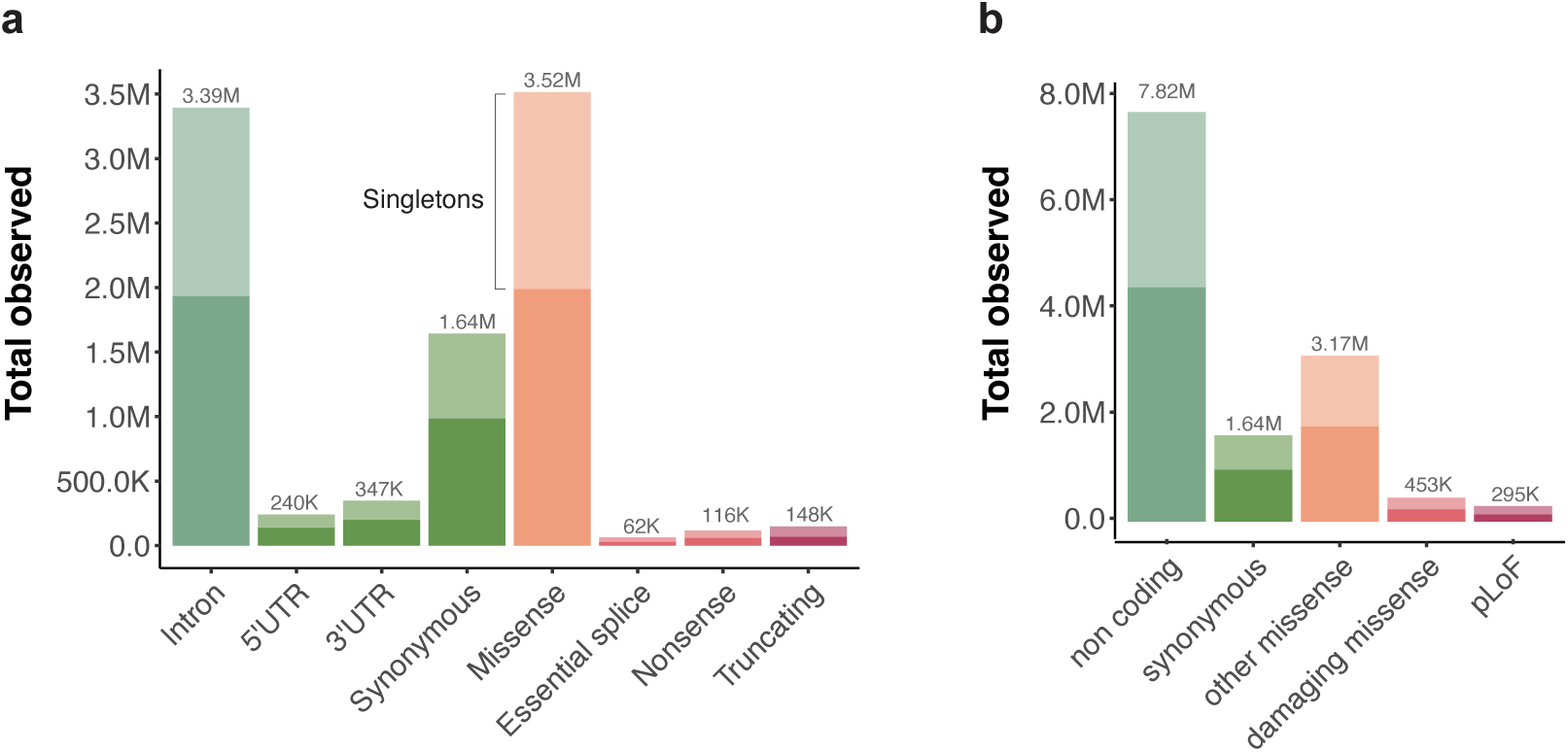
Distribution of variant annotation categories before and after broad consequence categorization. We annotate variants using VEP and by the most severe consequence in the canonical transcript. Panels (a) and (b) display the total number of unique variants observed across a set of variant consequences colored by degree of predicted impact, before and after broad variant consequence categorization. In each panel, green, orange and red colored bars indicate low, medium and high impact respectively, according to the color legends. Singleton variation within the variant class is stacked and displayed in a lighter shade. Counts of variant within each annotation category are displayed above the bars. Note that all counts shown here are before filtering to accurately phased variants.

**Supplementary Fig. 10:**
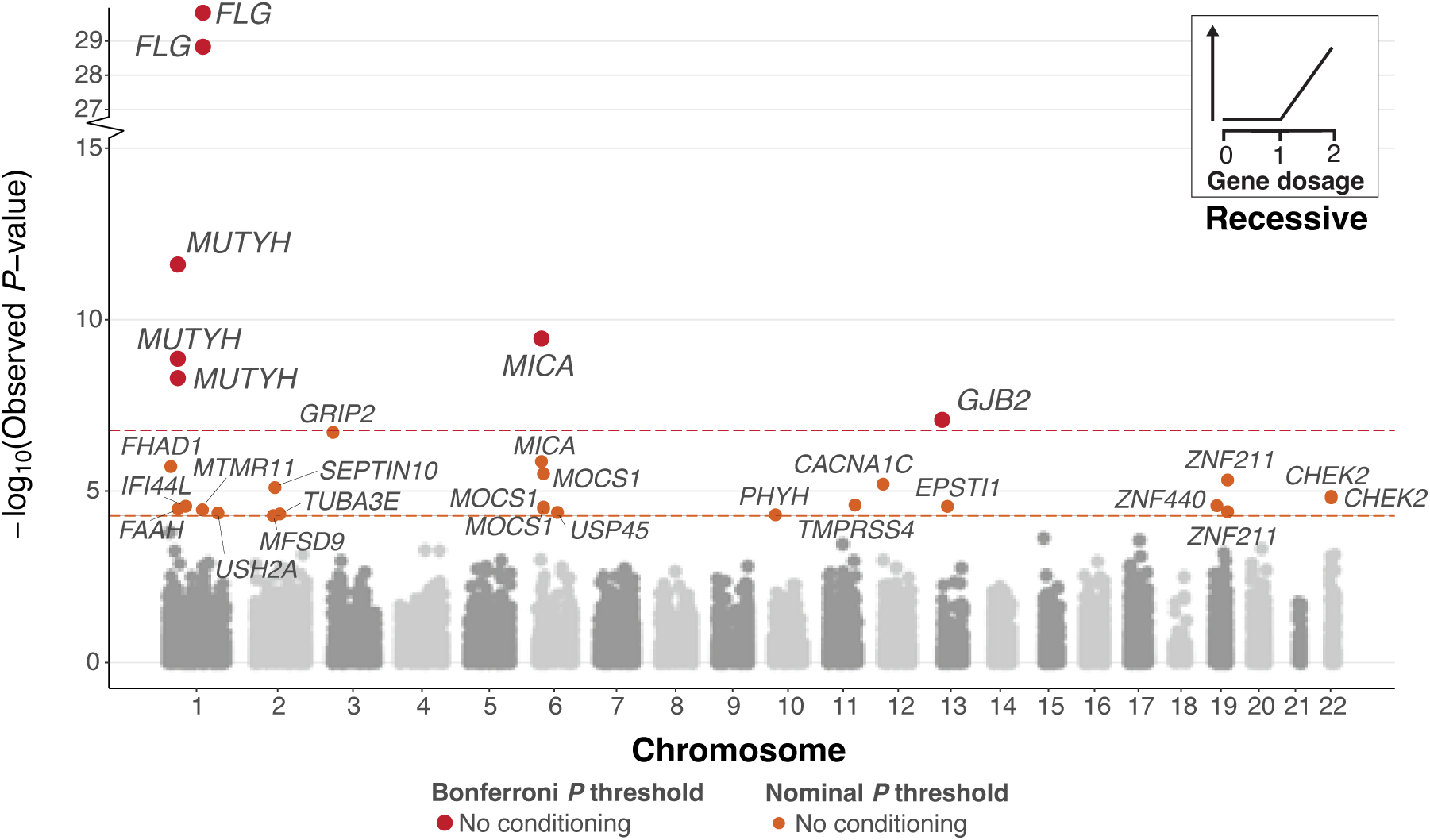
Recessive analysis of 311 phenotype without accounting for PRS. Recessive Manhattan plot depicting log_10_-transformed gene-trait association *P*-values versus chromosomal location. Associations are colored red or orange based on whether they are Bonferroni (𝑃 < 1.68 × 10^−7^) or nominally (𝑃 < 5.25 × 10^−5^) significant. No additional conditioning was carried out in this analysis.

### Gene copy dosage encoding and additive association modeling

We define annotate individuals as being ‘mono-allelic’ for a gene if they harbor at least one pLoFs or damaging missense variant with MAF < 5% on a single copy of the gene. Furthermore, if they harbor at least one pLoF or damaging missense variant on both inherited copies of the gene, we annotate them as ‘bi-allelic’. Using custom Hail scripts, we encode wildtypes, monoallelic and bi-allelic carriers as 0, 1 and 2 respectively, thus representing the number of affected gene copies in an individual. We test for association using SAIGE^63^, adjusting for sex, age, sex×age, age^2^, UKBB centre, genotyping batch and the first 10 PCs. Again, we took relatedness into account using a sparse GRM fitted on NFE. We restricted to gene-pairs with at least 10 disrupted haplotypes (corresponding to a minimum MAC ≥ 10), and adjust for multiple testing at Bonferroni significance (𝑃 < 0.05/gene-trait pairs).

**Supplementary Fig. 11:**
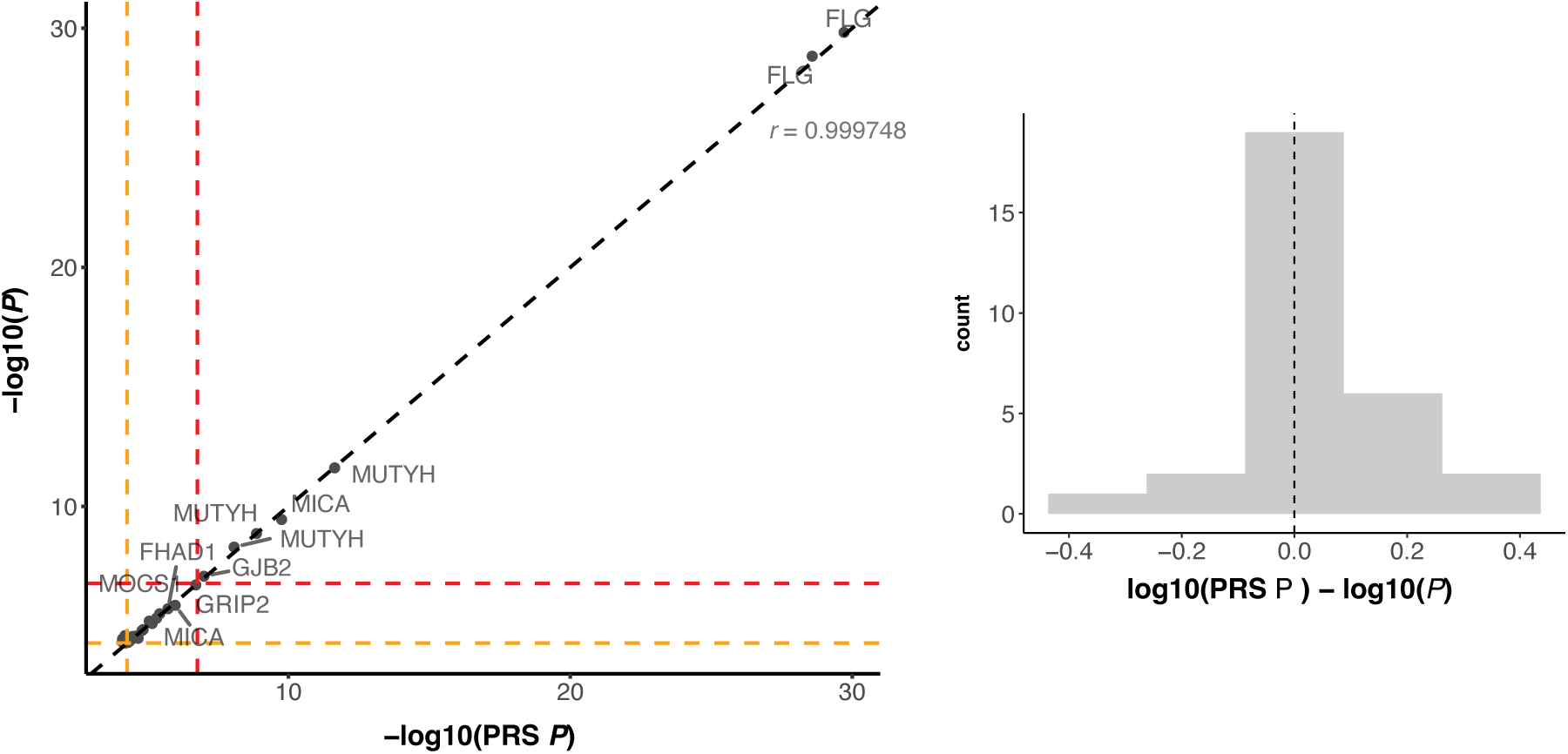
Association *P*-values before and after inclusion of PRS as a covariate. The scatter plot depicts the association P-values both before and after PRS was included as a covariate. The y-axis represents the P-value prior to PRS adjustment, while the x-axis demonstrates the P-value afterPRS adjustment. On the right, the difference in log-transformed P-values before and after PRS adjustment is displayed. The plot exclusively showcases gene-trait associations that were considered nominally significant in the recessive analysis.

## Polygenic risk scores

### Curation of array-based genetic data

We generated PRSs using imputed genotypes provided by UKBB^48^. In the following, we make the distinction between training and testing data. The first represents the samples that are used for fitting LDPred2^64^ weights and parameters while the latter represent the samples with bi-allelic variant (with homozygous or CH status) information in which we use to assess the predictive accuracy the fitted LDPred based PRS. For the training data, we took the genetically ascertained NFE and filtered to 246,152 unrelated samples (kinship coefficient < 2^−4.5^) that did not have quality controlled ES data available. NFE samples with high quality ES and imputed genotype data available were used for testing. Where predictive (nominal significant ℎ^2^ and 𝑛_𝑒 𝑓 𝑓_ ≥ 5000), we include PRS as a covariate for downstream biallelic association testing to account for common variant polygenic risk for the trait under investigation.

### Genotype variant filtering

We followed best practices from Privé *et al.*^64^, and filtered to common Haplotype Map 3 (HM3) SNPs^65^. Additionally, we exclude any variants with genotyping proportion < 1% and MAF < 1%, resulting in a total of 1,165,296 common autosomal variants for fitting PRS weights. To reduce the likelihood of spurious correlations between low-frequency variants in traits with low case or control count, we restricted to binary phenotypes with at least 1,250 cases and controls. Additionally, we imposed a phenotype specific MAF filter based on the number of cases and controls in a trait, specifically:

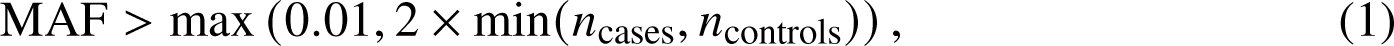

where 𝑛_cases_ and 𝑛_controls_ are the numbers of cases and controls with high quality imputed sequence data available, respectively, to guard against non-causal variants that are overrepresented in cases or controls leading to false positive associations.

### Common variant association testing

We tested for associations between the 1,165,296 common autosomal HM3 variants and phenotypes using Hail^46^, running logistic regression (logistic regression rows) adjusting for sex, age, sex × age, age^2^, UKBB assessment centre, genotyping batch and the first 10 PCs, using a Wald test.

### Estimating heritability

We generated LD-scores for HM3 variants in sample, using a random subset 10,000 of 246,152 unrelated genetically ascertained NFEs without haplotype information. Using the genome-wide association study (GWAS) summary statistics and LD-scores, we estimated SNP heritability *h*^2^_snp_ and standard errors (SEs) using LD score regression (LDSC)^66, 67^. We evaluated PRS for phenotypes with nominal significant *h*^2^_snp_ estimates (𝑃 < 0.05) and restricted to phenotypes with nominally significant (𝑃 < 0.05) LDSC based SNP heritability estimates and effective sample size 𝑛_eff_ ≥ 5, 000, where:

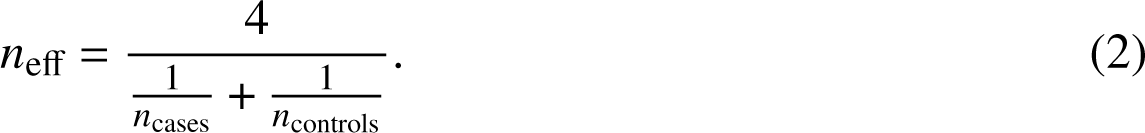

### Generating PRS using LDPred2

For a given phenotype, we trained a PRS predictor with LDPred2-auto^64^, using marginal effect size estimates evaluated on the 246,152 unrelated NFE samples (defined by kinship coefficient < 2^−4.5^) without ES data in the 200k ES UKBB release), *h*^2^_snp_ as estimated by LDSC, and in-sample reference panel to evaluate local LD, as input. We removed any invariant sites and mean-imputed missing genotypes, before training the predictor. Following PRS training, we then predict into the 176,266 samples with ES and high-quality imputed genotype data.

### Validation of polygenic risk scores

We assessed the ability of the resulting PRS to discriminate between case status by evaluating area under the curve (AUC) on the held-out unrelated set of samples with both HM3 SNPs and phased exome data. We used the function AUCBoot from the R package bigstatsr^68^ (1.5.6) to extract 10,000 bootstrap replicates of individuals and compute the 95% CIs for AUC.

## Conditional analysis

### Off-chromosome PRS conditional analysis

For each chromosome, 𝐶, we evaluated ‘off-chromosome’ PRS by setting weights on chromosome 𝐶 to 0. We repeated this for each phenotype with PRS available and fit SAIGE^63^ models while controlling for off-chromosome PRS by including it is as a covariate in the null SAIGE model.

### Common variant conditional analysis

To assess whether a putative signal in a gene is driven by nearby common variation, we filtered to samples that have both ES and imputed genotypes with MAF > 1% and imputation INFO score > 0.5. Then, for each gene that passed exome wide significance in the primary analysis (𝑃 < 5 × 10^−6^), we tested for common variant associations in the region (1 Mb upstream and downstream of the gene). For each of these regions, we took an iterative approach, testing for common variant associations using SAIGE^63^, conditioning on the lead variant and repeating the regression until the conditional 𝑃 for the newly included variant dropped below 5 × 10^−6^, allowing up to 25 ‘independent’ associations in the region. We used the same covariates as in the primary analysis. For every variant that passed exome-wide significance (𝑃 < 5 × 10^−6^), we encoded the genotypes as dosages and embedded them alongside pseudo variants (bi-allelic variants) in a VCF. We then re-ran the primary analysis twice (with and without controlling for off-chromosome PRS), while conditioning on any nearby common variant signals of association with the phenotype of interest.

### Rare and ultra-rare variant conditional analysis

For each significant (𝑃 < 1.68 × 10^−7^) gene-trait associations in the genome-wide analysis after conditioning on PRS and nearby common variants association signals, we considered a further conditioning step. We sought to determine whether the residual signal of association could be explained by additive rare variant effects within the associated gene. To do this, ran further conditioned on rare (MAC≥ 10, MAF≤ 0.05) and ultra-rare (MAC≤ 10) variants annotated as either pLoF or damaging missense within each gene. Because conditioning on ultra-rare variation can lead to convergence issues, we performed a gene-wide collapsing of ultra-rare (MAF≤ 10) variants, thus aggregating them into a single ‘super’ variant to represent burden of ultra-rare damaging variation in the gene. Following this collapsing, we were able to condition on the ultra rare and rare variant contribution using SAIGE, while also conditioning on PRS and nearby common variant association signals when applicable.

### Permutation of genetic phase

To test whether a putative gene-trait association is driven by compound heterozygosity, we designed a permutation-based pipeline that could be systematically applied and scaled across phenotypes and genes. To do this, we label samples that are either CH variants or heterozygous *cis* carriers and then randomly shuffle these labels a series of times. For each permutation, we re-run the association analysis conditioning on covariates as previously discussed (including off chromosome PRS and nearby common variants), and determine the resultant association strength under this label shuffling. Applying this permutation procedure multiple times, we can determine an empirical null for the association strength in the absence of phase information. The result is an empirical distribution of 𝑡-statistics and corresponding 𝑃-values that reflect the degree of association that would be expected given that the phase is random. We evaluate the one-sided empirical 𝑃-value, specifically:

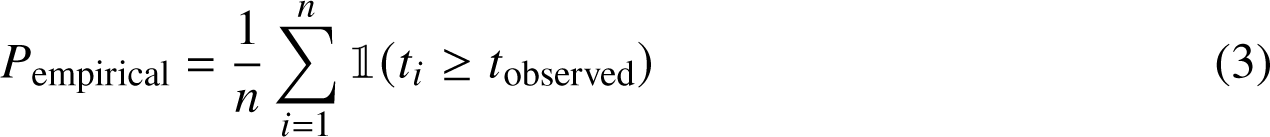

where 𝑛 is the number of permutations, 𝑡_𝑖_ is the 𝑡-statistic under the 𝑖^th^ random label shuffling, and 𝑡_observed_ is the observed 𝑡-statistic determined using the observed genetic phase. To ensure sampling of 𝑡-statistics at a sufficiently large number of configurations of the genetic phase, we analyzed gene-trait pairs with at least ten compound heterozygotes and/or samples with multiple variants on the same haplotype. We permuted up to 100,000 times. To control for multiple testing, we corrected for 5 gene-traits tested (Bonferroni significance threshold 𝑃 < 0.05/5 = 0.01).

## Gene-set enrichment of bi-allelic variation

### Analyzed gene-sets

We included the following gene lists in our gene-set enrichment analyses: essential in mice^69^, essential gnomAD^27^, essential ADaM^70^, essential in culture^71^, essential CRISPR^72^, genes with pLI > 0.9 in gnomAD^27^, non-essential in culture^71^, homozygous LoF tolerant^27^, and non-essential gnomAD^27^.

### Poisson regression to assess enrichment of CH variants in gene-sets

We test for depletion and gene-set enrichment using poisson regression. We model the count of bi-allelic variants across samples as a function of gene-set and mutation frequency using the glm function in R.

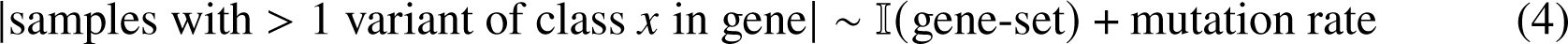

where 𝑥 is a pair (𝑥_1_, 𝑥_2_): 𝑥_1_ ∈ *{*pLoF, damaging missense, pLoF and/or damaging missense, other missense, synonymous*}*, 𝑥_2_ ∈ *{*heterozygote, CH, bi-allelic variants*}*. For each annotation category we use the transcript-specific mutation rate^28^. 95% confidence intervals are determined using confint.glm from the MASS-package (v7.3-58.1).

#### Homozygote and CH down-sampling

To investigate the number of identifiable CH or homozygous events across varying sample sizes and variant annotations, we performed down-sampling across the total population of 176,587 individuals. To do this, we defined a set of 35 regularly spaced cutoffs between 1,000 and 176,587 samples using increments of 5000. To determine uncertainty in our estimates of the number of unique genes implicated as a homozygote and/or CH, we randomly sampled individuals for each down-sampling 100 times, with replacement. We calculated the 95% CI by taking the 2.5% and 97.5% quantiles for the number of unique genes affected at a given sample size, and repeated across annotations (Fig. 19).

#### Power analysis for bi-allelic association

We perform a power analysis based on bi-allelic (including both CH and homozygous) variant frequencies in the population. To do this, we adopted code^73^ allowing us to determine the effective effect size on the OR scale across candidate configurations of binary case-control counts by substituting alternate allele frequencies with bi-allelic variant frequencies. We calculated effect sizes at 80% power at Bonferroni significance (𝑃 < 1.7 × 10^7^) for a hypothetical traits with 823 (0.5%), 1766 (1%), 3532 (2%), 5298 (3%), 8829 (5%) cases of 176,587 total samples.

**Supplementary Fig. 12:**
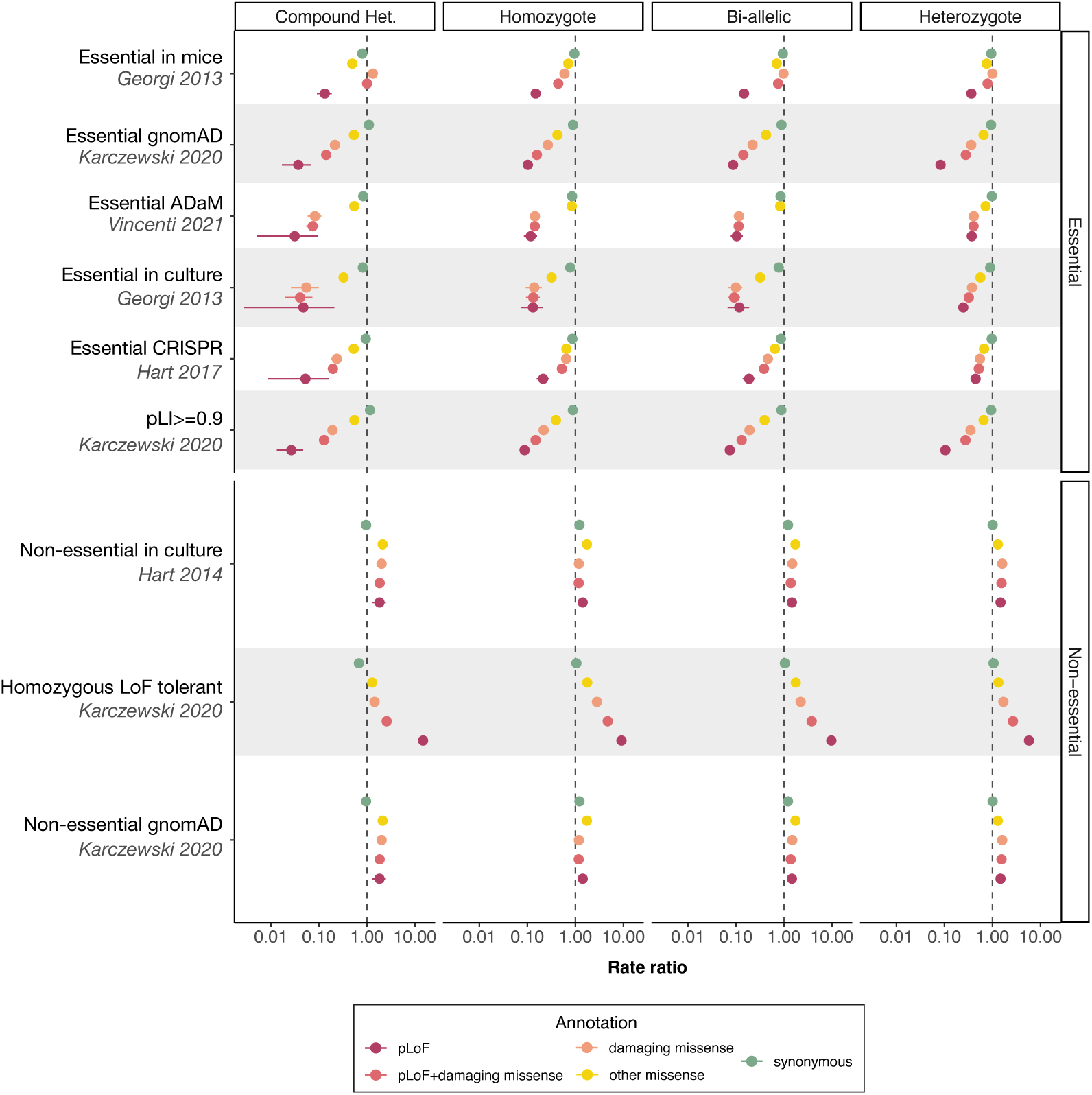
Gene-set depletion/enrichment modeling. Poisson regression to model mono- and bi-allelic variant (heterozygous, CH, homozygous or both) depletion and enrichment across essential and non-essential gene-sets. Rate ratios are shown for synonymous (green), other missense (yellow), damaging missense (orange) and pLoF (red) variants. The dashed line depicts a rate ratio of 1.

**Supplementary Fig. 13:**
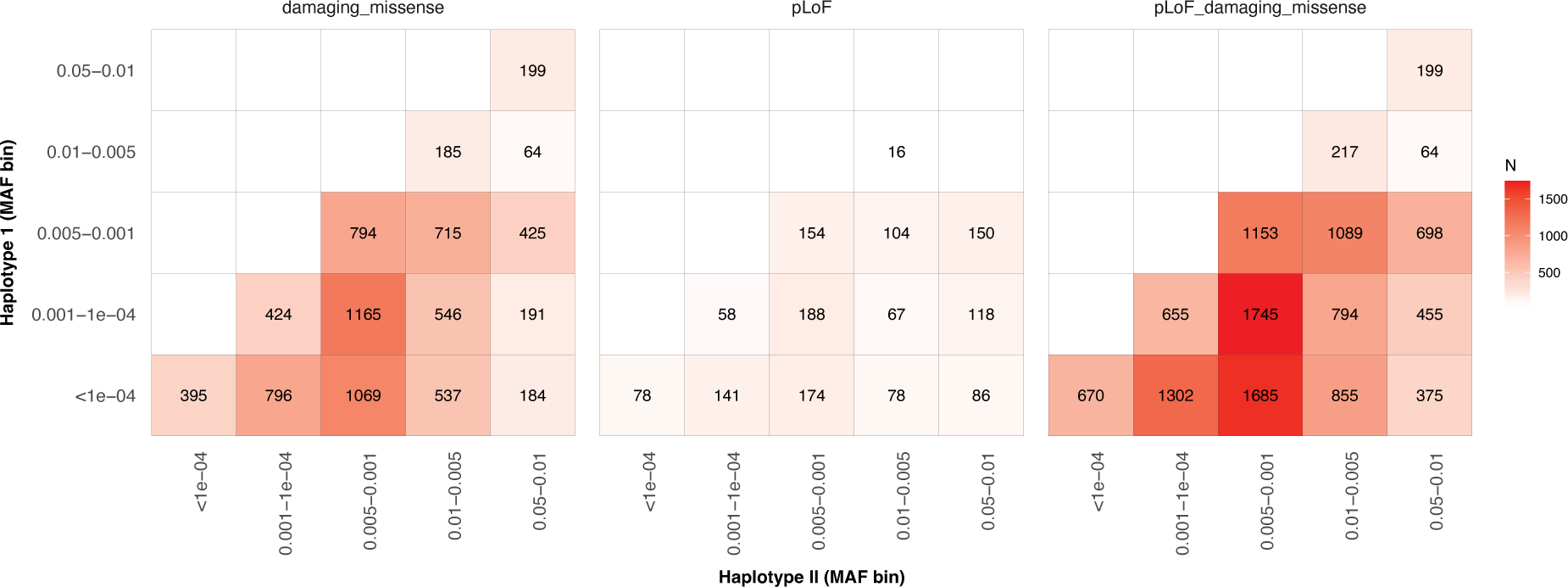
Allele frequencies of variants in the CH state. Heatmap of allele counts for variants in CH state stratified by predicted variant consequence (damaging missense, pLoF or pLoF+damaging missense). We plot the MAC for variants residing on the most common haplotype (y-axis) versus the rarest haplotype (x-axis).

**Supplementary Fig. 14:**
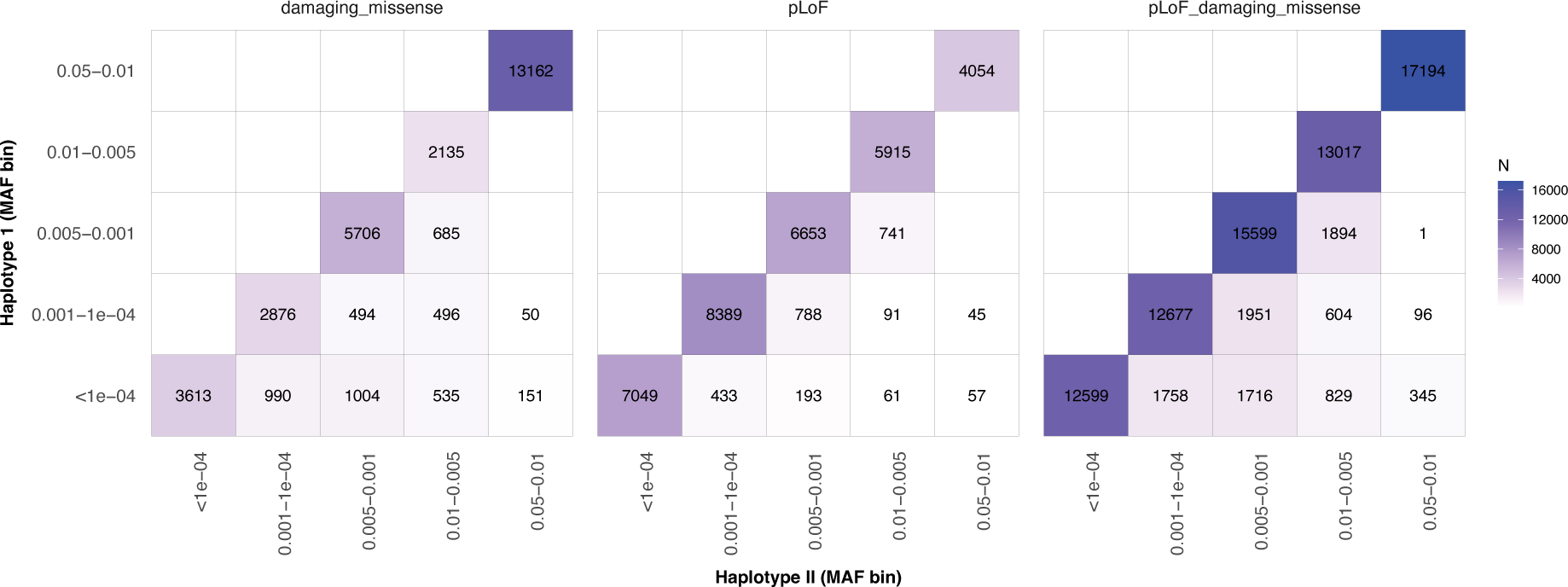
Allele frequencies of variants in *cis*. Heatmap of allele counts for co-occurring variants on the same haplotype stratified by predicted variant consequence (damaging missense, pLoF or pLoF+damaging missense). The most common variant on the haplotype versus the least common are plotted.

**Supplementary Fig. 15:**
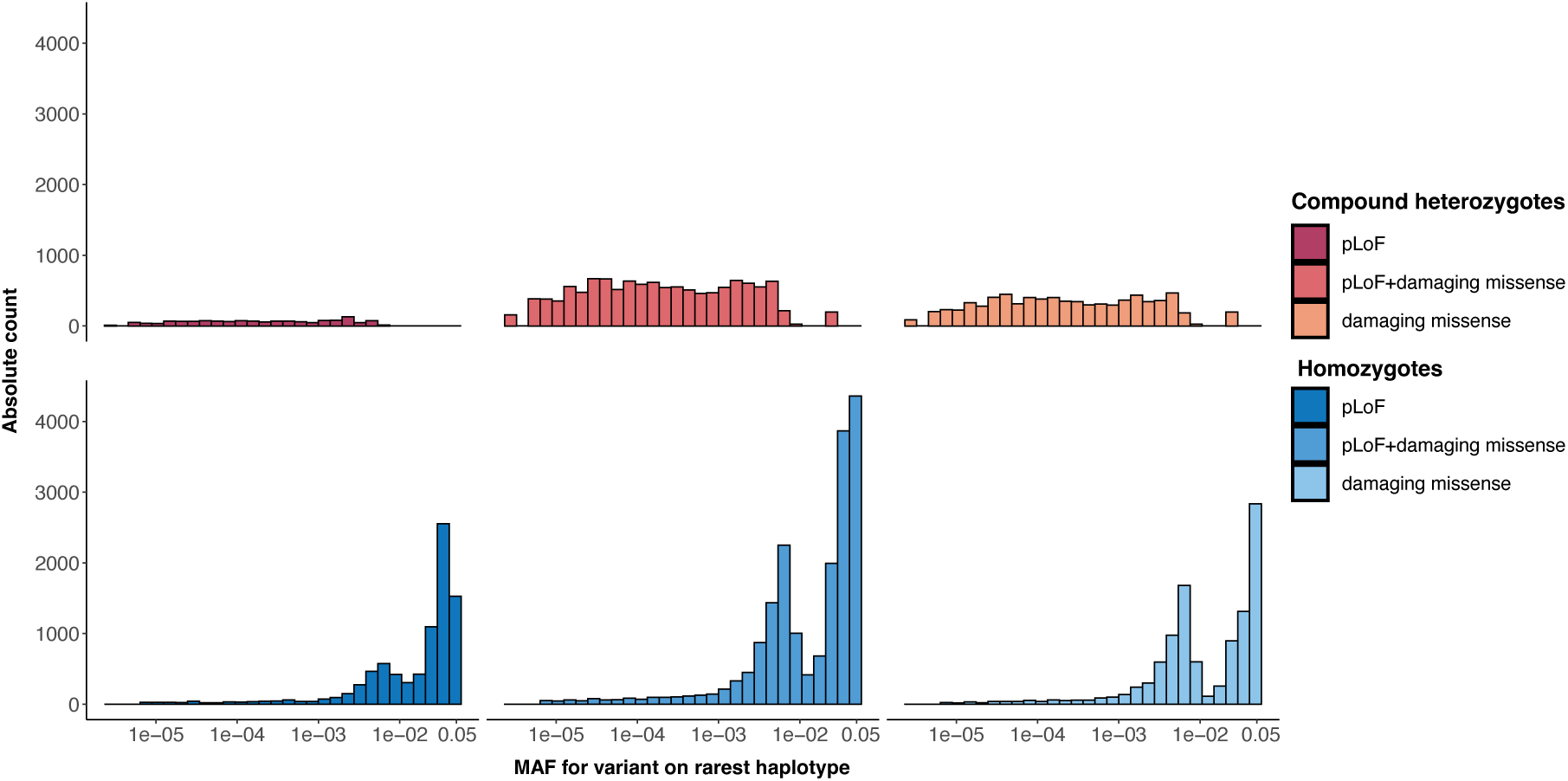
Distribution of observed variants across samples by allele frequency. Histogram of unique bi-allelic variant (CH and homozygotes) prevalence across the allele frequency spectrum. For a qualifying CH variant, the allele frequency corresponding to the alternate allele on the rarest haplotype are plotted.

**Supplementary Fig. 16:**
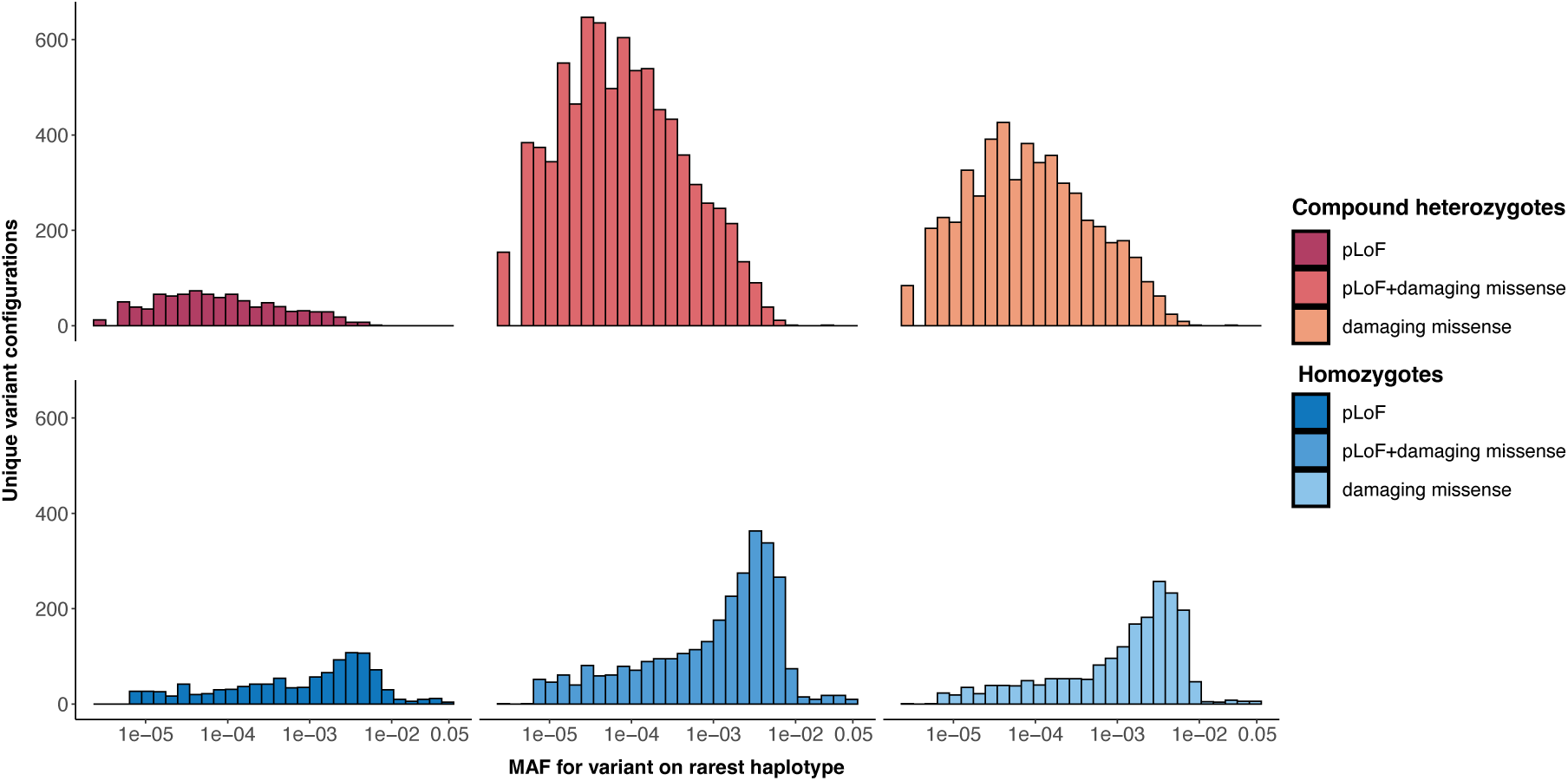
Distribution of unique variants observed by allele frequency. Histogram of bi-allelic variant (CH and homozygotes) count for all gene-samples pairs in the analysis. For a qualifying CH variant, the allele frequency corresponding to the alternate allele on the rarest haplotype are plotted.

**Supplementary Fig. 17:**
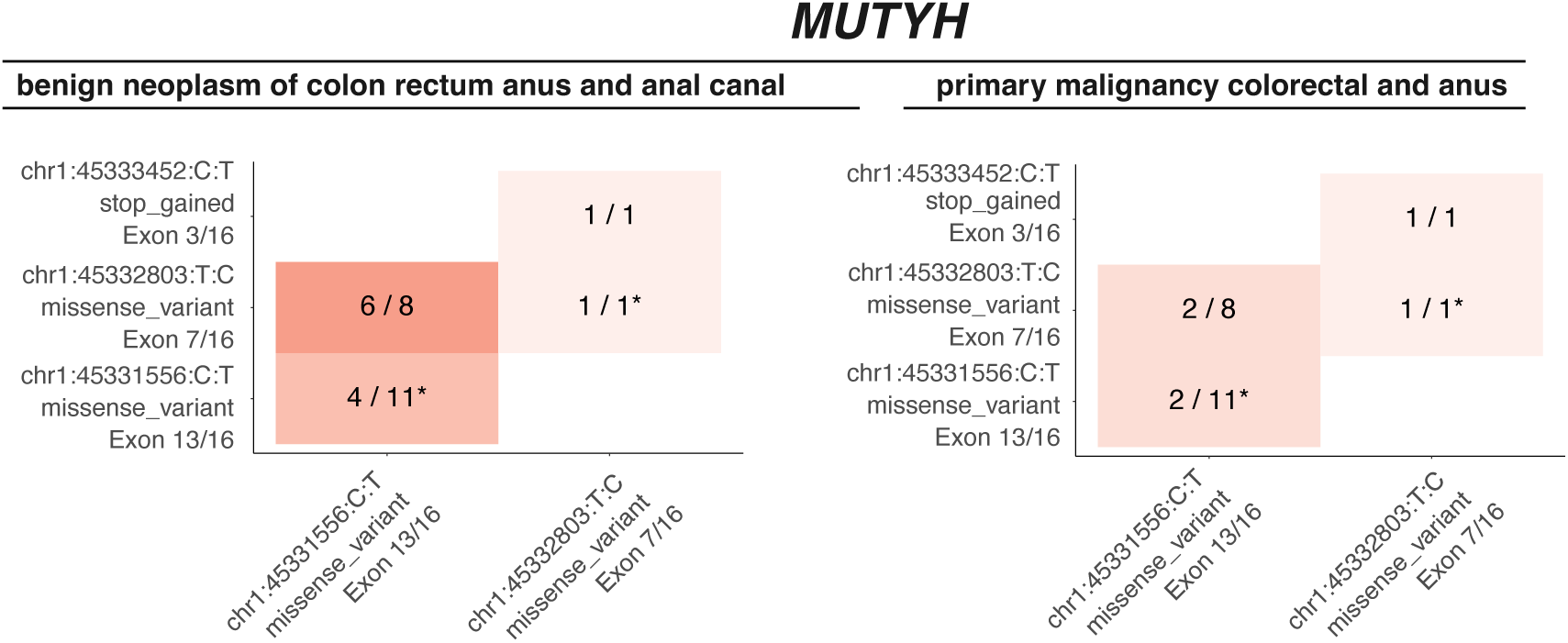
Co-occurrence of deleterious MUTYH-variants across colorectal cancer outcomes. Bi-allelic variant occurrence in *MUTYH* for benign neoplasm of the colon, rectum and anal canal (left) and primary malignancy of the colon (right). The constituent variants are shown alongside the variant consequence and involved exon or intron. Each cell indicates that number of individuals that are cases out of the total bi-allelic carriers identified. Homozygous cases and carriers are indicated with a star (*)

**Supplementary Fig. 18:**
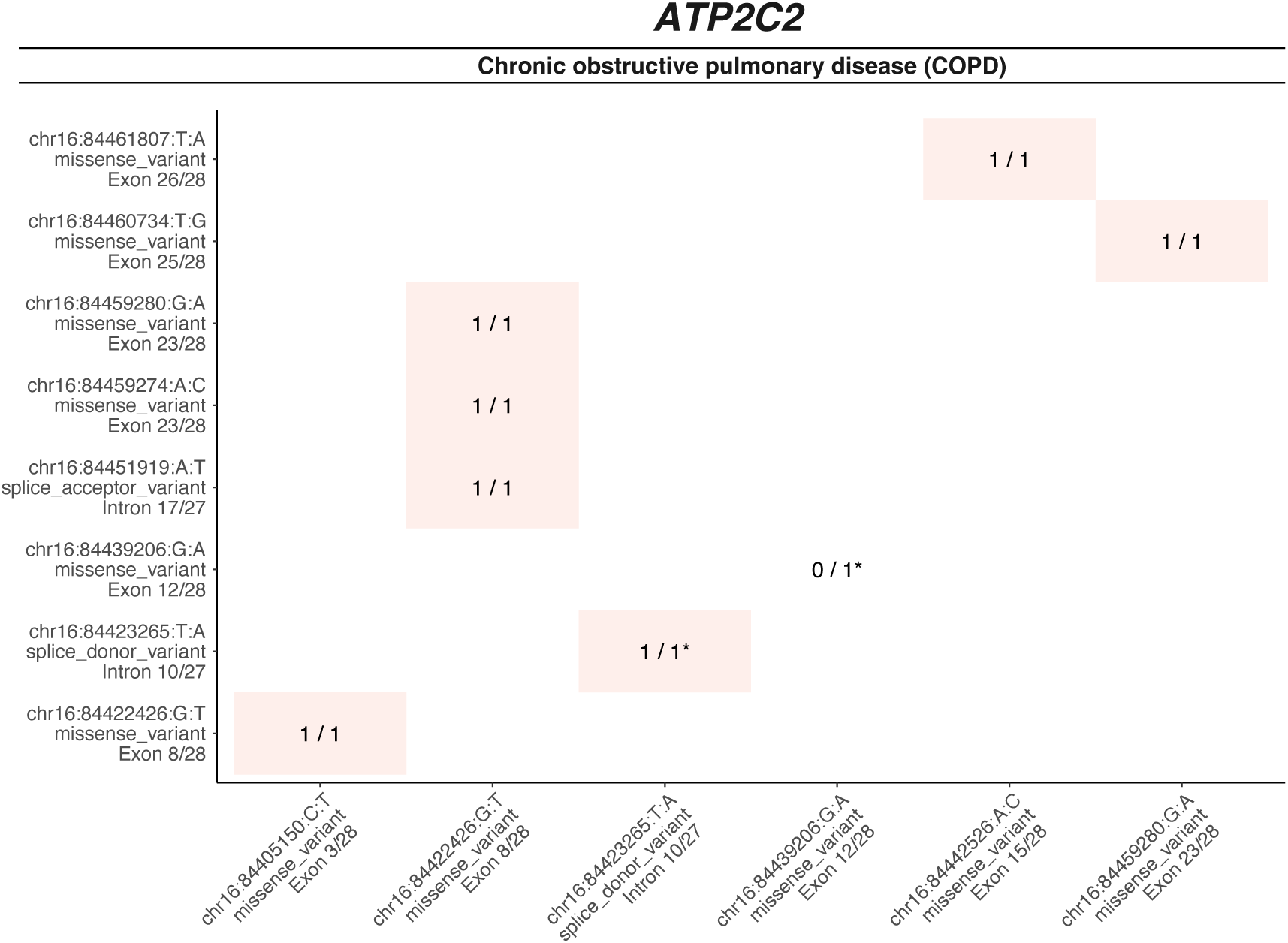
Co-occurrence of deleterious ATP2C2 variants by COPD status. Bi-allelic variant occurrence in *ATP2C2* for chronic obstructive pulmonary disease (COPD). The constituent variants are shown alongside the variant consequence and involved exon or intron. Each tile indicates that number of individuals are cases out of the total bi-allelic carriers identified. Only the variants that affect both gene copies are shown. Stars (*) are included in the label to indicate homozygosity.

**Supplementary Fig. 19:**
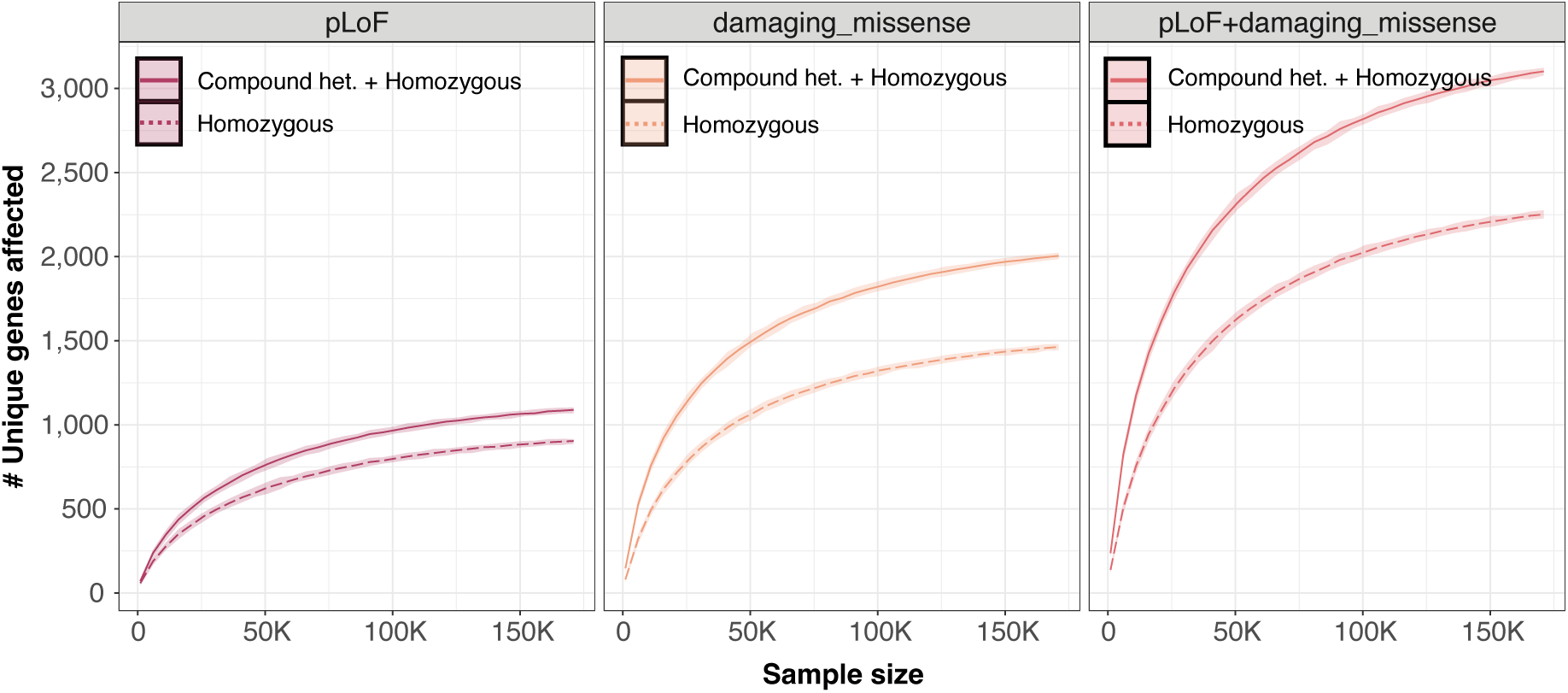
Count of unique genes affected by a homozygous and homozygous or compound heterozygous variants as a function of sample size. Starting with the full data, and down-sampling, we plot counts number of unique genes harboring homozygous and homozygous or compound heterozygous variants as a sample size is decreased. Class of variants in each count are denoted according to the key. Each facet indicate a specific variant annotation.

## Simulation

### Simulation of synthetic phenotypes using real genotypes

We performed a series of simulations to test that our pipeline would detect a CH effect in the presence of a true signal. We sampled 100,000 genetically-ascertained NFEs in the UKBB data, and extract chromosome 22 which we then use to simulate phenotypic data with a recessive genetic architecture. To emulate a scenario in which defects in protein coding genes lead to disease, we annotated the filtered UKBB genetic data and determined the collection of samples harboring damaging bi-allelic variants in each gene (compound heterozygous and homozygous, comprised of variants annotated as pLoF or damaging missense). We then define a 𝑛 samples × ^B^ 𝑚 genes matrix **B̃** with entries:

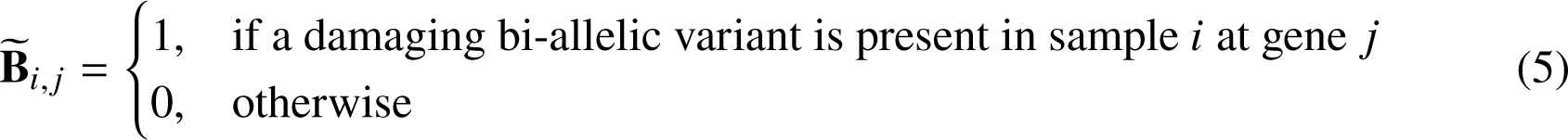

We then simulated liability under the following model:

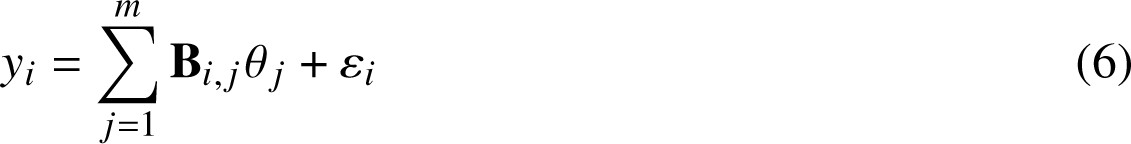

where B_𝑖, 𝑗_ is the (𝑖, 𝑗)^th^ entry of B after standardizing the columns of 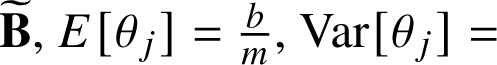 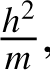. Here, we implicitly assume that presence of at least one homozygous or CH variant of any type within a given gene contributes the same risk to disease, whose average across genes is set by the parameter 𝑏. The resultant liability 𝑦_𝑖_ has mean 0 and variance 1. Note that the standardization of B imposes a frequency dependent relationship between prevalence of bi-allelic damaging variants in a gene and variance explained. We simulated under the spike-and-slab model:

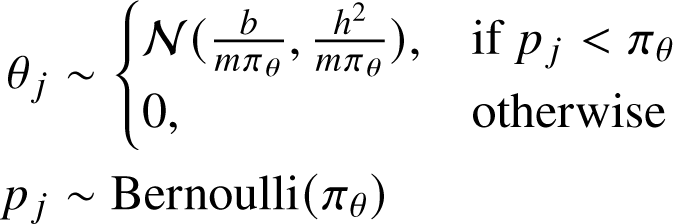

in which 𝜋_𝜃_ ∈ [0, 1] is the proportion of causal genes with a recessive contribution to the phenotype. Finally, to obtain binary traits we used the liability threshold model assuming a case prevalence of 10%. In the following simulations, we set 𝜋_𝜃_ = 0.25, and considered ℎ^2^ values of ℎ^2^ ∈ {0, 0.01, 0.02, 0.05, 0.10} and 𝑏 values of 𝑏 ∈ {0, 0.5, 1, 2, 10}.

**Supplementary Fig. 20:**
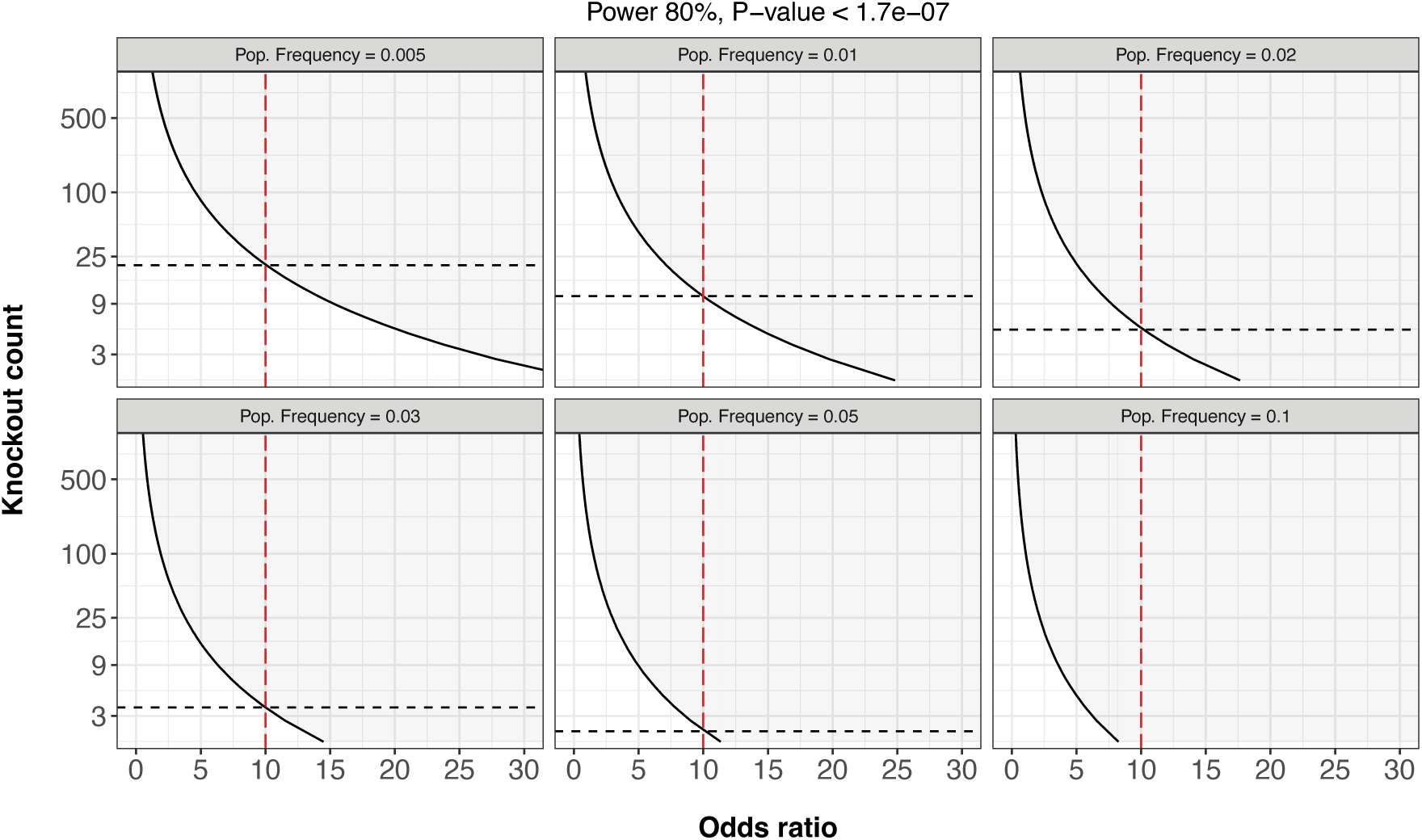
Power analysis to determine the required number of bi-allelic variants to detect specific ORs at 80% power at bonferroni significance (𝑃 < 1.7 × 10−7). We repeat the analysis while varying trait population prevalence assuming 823 (0.5%), 1766 (1%), 3532 (2%), 5298 (3%), 8829 (5%) cases out of 176,587 total individuals. The dashed red lines in the plot demonstrate the required number of bi-allelic variants to detect an OR ≥ 10.

**Supplementary Fig. 21.**
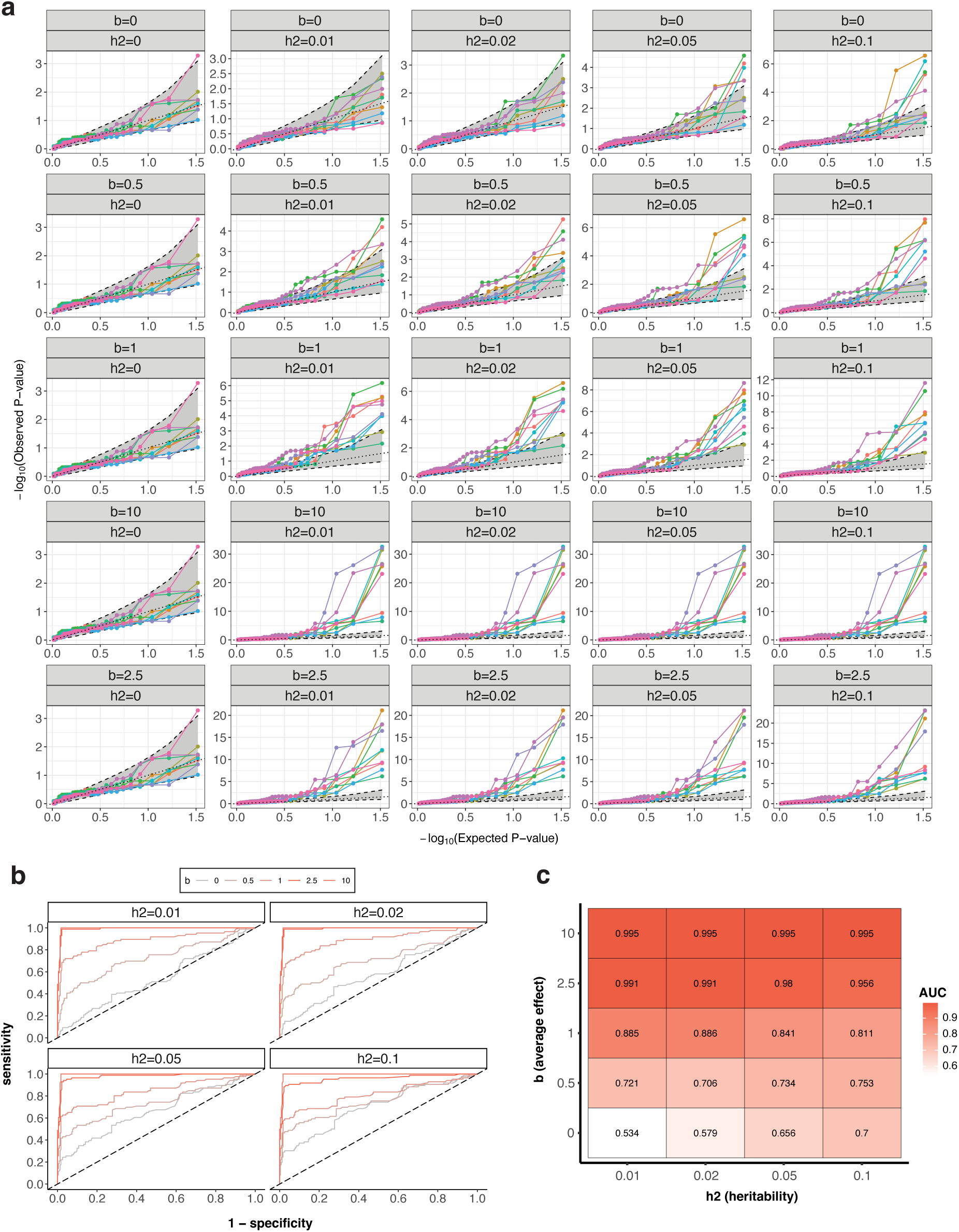
*(previous page)*: Simulation study to test our ability to detect biallelic effects in the presence of true effects. We simulate phenotypic data applied to 100,000 genetically-ascertained NFE on chromosome 22 (Methods) under the liability-threshold model assuming a spike and slab genetic architecture. We assume a 10% disease prevalence and 25% causal genes, and consider varying levels of phenotypic variance explained by these effects ∈ {0, 0.01, 0.02, 0.05, 0.10}. We then apply SAIGE to the simulated phenotypes, testing for an association between presence of a bi-allelic variant in each gene and case status. **a)** Each panel indicates a set of simulations assuming varying levels of heritability and average effect as labeled in the subtitles. In each panel, we plot the true effect size in the simulation for a given gene on 𝑥-axis against the corresponding − log_10_(𝑃) value of association. Areas of circles correspond to the number of samples harboring bi-allelic damaging variants in the 100,000 samples according to the legend. **b)** To assess the sensitivity and specificity of our approach, we created ROC-AUC curves for each combination of increasing phenotypic variance explained (facet) and increasing average affect (red lines). **c)** For each ROC-AUC curve from b, we calculate the AUC. White indicates low AUC and red indicates higher AUC.

## Longitudinal effects

### Time-to-event data curation

We curated age-at-diagnosis for 278 binary phenotypes from the UKBB-linked primary care and hospital record data. 251 phenotypes were curated using the mapping tables generated by Kuan *et al.*^55^, excluding any codes related to “history of…” events for which accurate age-at-diagnosis could not be extracted. The remaining 27 phenotypes individuals’ records were left-truncated at the age of first record (of any code) in either the primary care or hospital data, and right-censored at the age of the last record.

### Cox proportional-hazards modeling

For each gene-trait combination to test, we performed Cox-proportional hazards modeling to estimate differences in lifetime risk of developing the phenotype between heterozygous carriers of pLoF + damaging missense variants in the gene (reference group) and individuals who are bi-allelic carriers (compound-heterozygous or homozygous), multi-hit *cis*-heterozygous carriers, and wildtypes. All effects were adjusted for sex, the first 10 genetic PCs, birth cohort (in ten-year intervals from 1930-1970), and UKBB assessment center. For phenotypes with a significantly heritable PRS, we additionally adjusted for off-chromosome PRS. We visualized survival probabilities using Kaplan-Meier curves^74^. Finally, for gene-trait combinations where we were powered to detect differences between compound-heterozygous and multi-hit *cis* heterozygous carriers of variants, i.e. where each group contained at least five cases of the phenotype, we repeated the above analysis with multi-hit *cis* heterozygous carriers as the reference group. Cox proportional-hazards regression was performed using the R package survival 3.3.1^75^ and Kaplan-Meier plots drawn with the R package survminer 0.4.9^76^.

**Supplementary Fig. 22:**
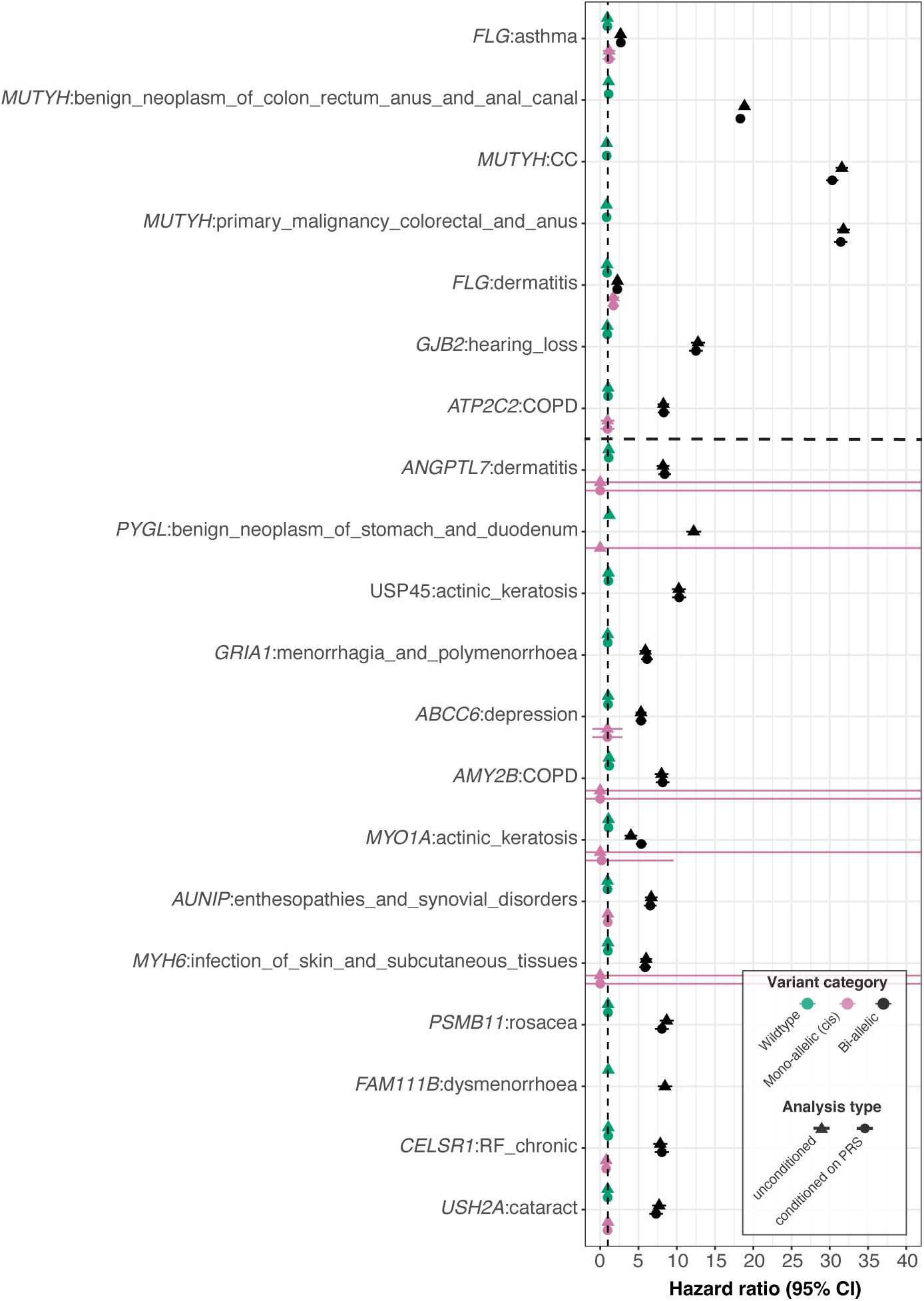
Cox proportional hazards modeling with and without polygenic effects. HRs when comparing CH and homozygous status versus heterozygous carrier status. Throughout, we display hazard ratios with (circles) and without (triangles) taking the polygenic contribution into account by conditioning on off-chromosome PRSs for heritable traits that pass our quality control cutoffs. HRs for gene-traits with one or more individuals with multiple cis variants on the same haplotype are also displayed in pink. Associations that pass Bonferroni significance (𝑃 < 1.89 × 10^−7^) and FDRs < 5% cutoff are demarcated by the dashed line in the top and bottom half respectively. Abbreviations: CC (colorectal cancer), COPD (chronic obstructive pulmonary disease). ^71^

**Supplementary Fig. 23:**
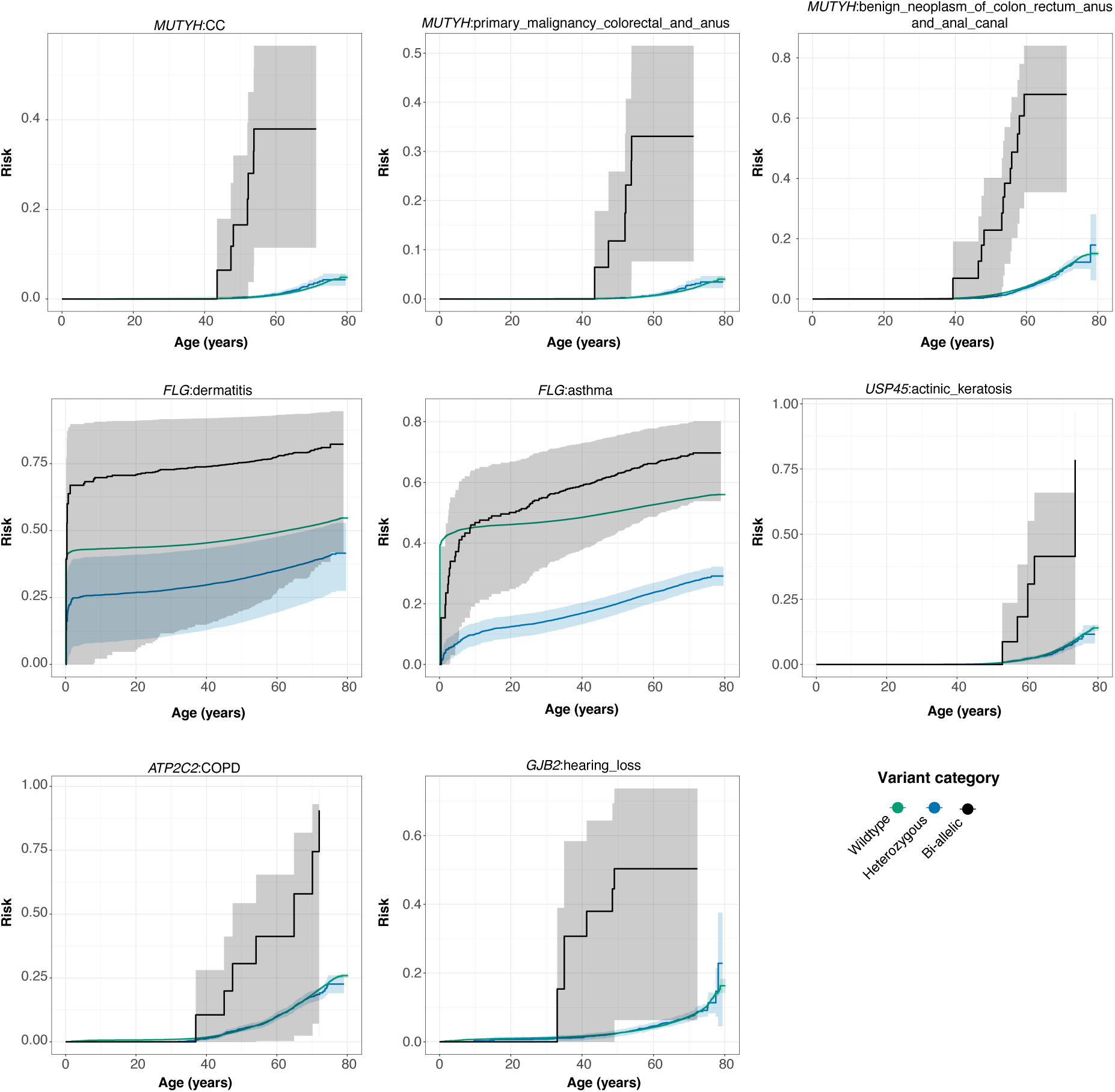
Kaplan-Meier survival curves for carriers of bi-allelic variants. Trajectories for wildtypes and bi-allelic (CH or homozygous) carriers of damaging missense/protein-altering mutations are shown with green and black lines respectively. For traits where over 50% of cases are left-censored, the confidence interval estimates cannot be accurately determined using Kaplan-Meier curves, and thus, these should be disregarded. Consequently, wildtype confidence intervals for *FLG*-Asthma and *FLG*-Dermatitis are not displayed in the figure.

**Supplementary Fig. 24:**
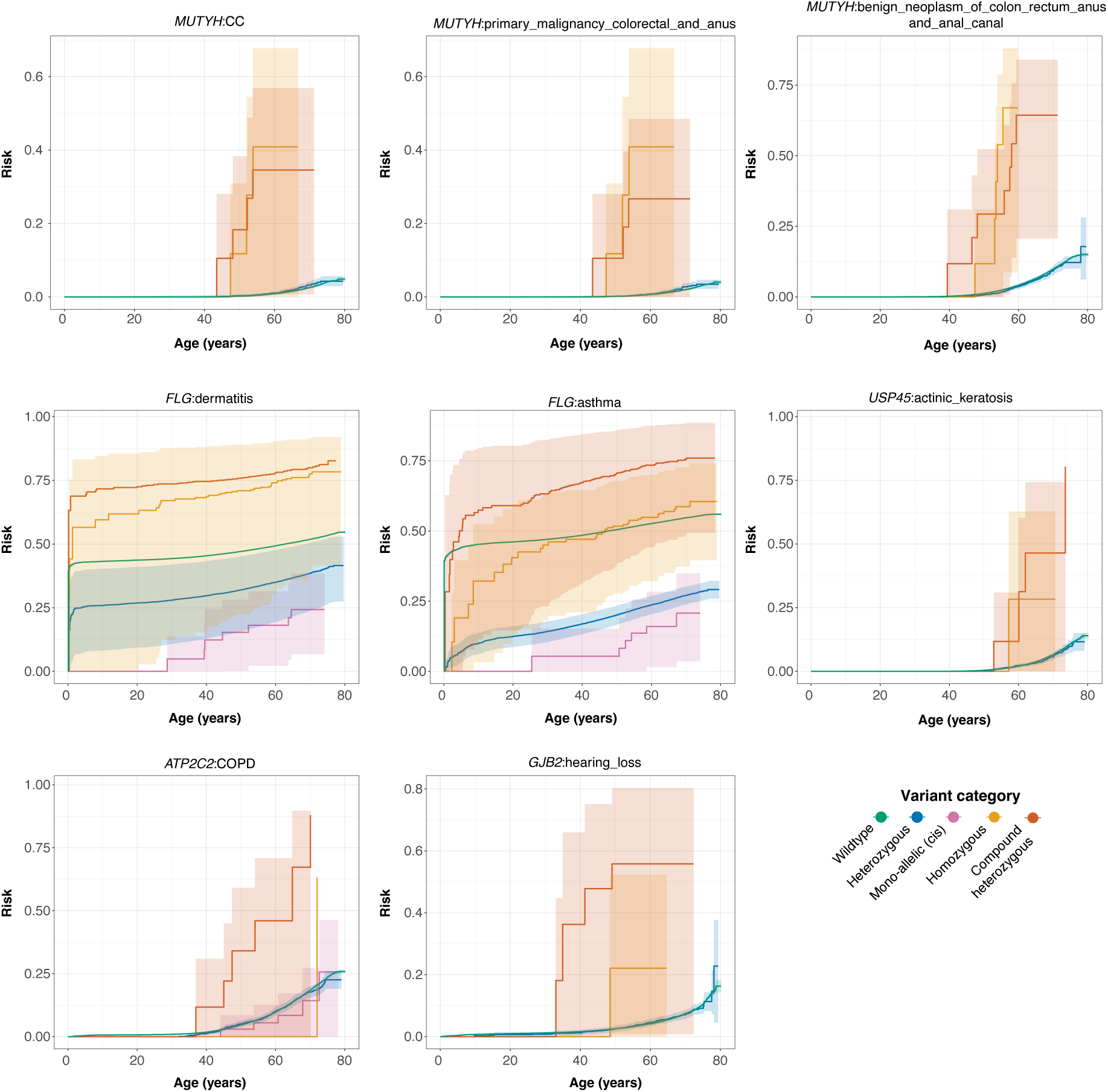
Kaplan-Meier survival curves for carriers of CH, homozygous, heterozygous variants. Kaplan-Meier survival curves for CH (red), homozygous (orange), heterozygous carriers (blue), single disruption of haplotypes (pink) owed to pLoF or damaging missense/protein-altering mutations. Wildtypes are shown in green. For traits where over 50% of cases are left-censored, the confidence interval estimates cannot be accurately determined using Kaplan-Meier curves, and thus, these should be disregarded. For this reason, wildtype confidence intervals for *FLG*-Asthma are not displayed in the figure. Wildtype and CH confidence intervals are also not shown for *FLG*-Dermatitis.

## Supplementary Tables

**Supplementary Table 4: Comparative analysis of SER point estimates for phasing Methods: SHAPEIT4, SHAPEIT5 (pre and post-filtering by phasing confidence), and Eagle2.** This table presents a comparison of the SER point estimates for various phasing methods, including SHAPEIT4, SHAPEIT5 (both before and after filtering by phasing confidence), and Eagle2. It is important to note that we employed the full phased set of autosomes for SHAPEIT5, while for SHAPEIT4 and Eagle2, we limited our analysis to chromosomes 20, 21, and 22. The table presents 95% confidence intervals for each method.

**Supplementary Table 5: Trio-SER across final reconstituted chromosomes for variants originating from ES and genotyping array data before and after filtering by PP ≥ 0.90.** All confidence intervals are 95% binomial confidence intervals.

**Supplementary Table 6: Trio-SER by MAC bin for 96 parent-offspring trio relationships before and after filtering by PP ≥ 0.90.** All confidence intervals represent the 95% binomial confidence interval.

**Supplementary Table 7: Number of trio-switch errors binned by genes before and after filtering by PP** ≥ 0.90 using 93 trio-offspring relationships.

**Supplementary Table 8: Comparative analysis of predicted phase Using SHAPEIT5 vs. read-backed phasing with Whatshap, subsetting by PP** ≥ 0.90. This comparison focuses on the predicted phase generated using SHAPEIT5 after subsetting by PP ≥ 0.90 against the read-backed phased variants determined through Whatshap. The comparison is limited to pairs of variants within a read. The analysis was conducted across chromosomes 20, 21, and 22, utilizing approximately 176,000 genetically ascertained NFE samples. In the ‘Errors’ column, errors are defined as discrepancies between the genetic phase of statistically phased variants (processed with SHAPEIT5) and read-backed phased variants (determined with Whatshap). Binomial confidence intervals are used throughout.

**Supplementary Table 9: Annotation of the most severe consequence for variants in canonical transcripts across 22 autosomes for quality-controlled variants and samples, pre- and post-filtering to PP** ≥ 0.90. The table enumerates the most severe predicted consequences of variants located in canonical transcripts, analyzed across 22 autosomes for variants and samples that underwent stringent quality control, both before and after filtering to PP ≥ 0.90 .

**Supplementary Table 10: Results of the Poisson regression analysis used to evaluate the enrichment (rate ratios) of both mono-and bi-allelic variants within gene-sets.** We applied this model to assess both the depletion and enrichment of gene-sets using Poisson regression. The count of bi-allelic variants across samples is modeled as a function of the gene-set and mutation frequency.

**Supplementary Table 11: Tally of predicted carriers among 176k individuals classified as CH, homozygous, on the same haplotypes (*cis*), or bi-allelic (either CH and/or homozygous)**

**Supplementary Table 12: Estimation of heritability and computation of polygenic risk by applying LDSC and LDPred2 to HapMap3 SNPs to a dataset of 246k samples without phase information.** This is followed by prediction of polygenic risk for each individual within a subset of 176k phased samples. PRS that satisfy our filtering criteria (based on LDSC 𝑃-value and effective sample size) are subsequently incorporated (Methods). Evaluation of accuracy is performed through a non-parametric bootstrap method, involving calculation of the AUC and its associated standard errors.

**Supplementary Table 13: Systematic search and conditional analysis of common variants around significant gene-trait associations.** This table presents the results of our systematic search for common variants (MAF > 1%) located within 1 MB upstream and downstream of any significant gene-trait associations (𝑃 < 5.25 × 10^−5^). Upon identifying a significant common variant (𝑃 < 5 × 10^−6^), a conditional analysis was performed using that variant. If other significant variants remained, they were included in subsequent iterations of the analysis until either the signal was exhausted, or 25 iterations were completed. The table enumerates all the resulting gene-traits and variants upon which we have conditioned.

**Supplementary Table 14: Overview of significant results obtained before and after conditioning on off-chromosome PRS, nearby common variation, and the burden of rare variation.** The analysis was also carried out for compound heterozygotes and homozygotes independently while conditioning on PRS when applicable (methods). In addition, we aggregated rare variants by haplotype and modeled the number of affected haplotypes in each individual. Subsequently, we performed two analyses: (1) an additive association analysis using the haplotype encoding, and (2) a recessive association analysis, conditioned on the additive encoding of haplotypes.

**Supplementary Table 15: Overview of nominally significant (𝑃 < 3.05 × 10^−6^) and Bonferroni-corrected significant hits (𝑃 < 9.8 × 10^−9^) hits from additive association analysis by modeling the number of putatively disrupted haplotypes per individual.** We restrict to genes in which there are at least 10 total disrupted haplotypes in the population. Throughout the analysis, we condition on off-chromosome PRS when applicable.

**Supplementary Table 16: Significant associations (FDR<5%) in Cox proportional-hazards models when comparing compound heterozygous and homozygous carriers against heterozygous carrier status.** We take the polygenic contribution into account by conditioning on off-chromosome PRS for heritable traits that pass our quality control cutoffs (Methods).

**Supplementary Table 17: Cox proportional-hazards ratios when comparing compound heterozygous and homozygous multiple variants on the same haplotype and wildtype status versus heterozygous carrier status.** We take the polygenic contribution into account by conditioning on off-chromosome PRS for heritable traits that pass our quality control cutoffs (Methods).

**Supplementary Table 18: Cox proportional-hazards ratios when comparing compound heterozygous and homozygous, heterozygous and wildtype status against carriers of multiple variants on the same haplotype.** We take the polygenic contribution into account by conditioning on off-chromosome PRS for heritable traits that pass our quality control cutoffs (Methods).

**Supplementary Table 19: Median age of diagnosis across for pLoF+damaging missense carriers that are heterozygous, homozygous, compound heterozygous, bi-allelic (homozygous and/or compound heterozygous) and have multiple variants on the same haplotype (*cis*).**

**Supplementary Table 20: Overview of variants co-occurring in significant (FDR < 5%) Cox-proportional hazards gene-trait combinations**

